# Mendelian Randomization analyses identify causal associations of human gut microbiome composition on intelligence

**DOI:** 10.1101/2023.05.11.23289760

**Authors:** Shi Yao, Ji-Zhou Han, Xin Wang, Jia-Hao Wang, Long Qian, Hao Wu, Jing Guo, Shan-Shan Dong, Yan Guo, Tie-Lin Yang

## Abstract

**Background:** Growing evidence indicates that dynamic changes in the gut microbiome can affect intelligence; however, whether the relationships are causal is unknown.

**Methods:** We conducted a bidirectional two-sample Mendelian randomization (MR) analysis using the summary statistics from the largest GWAS meta-analysis of gut microbiota composition (n = 18,340) and intelligence (n = 269,867). Inverse-variance weighted method was used to conduct the MR analyses complemented by a range of sensitivity analyses to validate the robustness of the results. We further applied a two-step MR analysis to evaluate whether the effect of identified taxa on intelligence was mediated by regulating the brain volume.

**Results:** MR evidence suggested a risk effect of the genus *Oxalobacter* on intelligence (β = –0.032; 95% confidence interval, –0.049 to –0.015; *P* = 1.88 ×10^-4^) and a protective effect of the genus *Fusicatenibacter* on intelligence (β = 0.051; 95% confidence interval, 0.023 to 0.079; *P* = 3.03× 10^-4^). In the other direction, we did not find causal evidence of intelligence on gut microbiome composition. The mediation analysis showed that the effect of genus *Fusicatenibacter* on intelligence was partly mediated by regulating the brain volume, with a mediated proportion of 26.7% (95% confidence interval, 4.9% to 48.5%).

**Conclusions:** Our findings may help reshape our understanding of the microbiota-gut-brain axis and development of novel intervention approaches for preventing cognitive impairment.

## Introduction

Intelligence, also known as cognitive ability, is a robust predictor of educational and socioeconomic achievement and broadly implies lifestyle behaviors and health resource advantages across the lifespan [1–3]. Establishing causality and prioritizing targets responsible for individual differences in intelligence is one of the key challenges in psychological and brain sciences. Currently, emerging evidence recognizes gut microbiota as an essential component of normal physiology, with an important role in both brain development and function [4–6].

The gut microbiome is a highly complex and diverse hidden kingdom and plays a fundamental role in gut-brain communication. Growing evidence indicates that alterations in the gut microbiome can affect neurodevelopment and cognitive ability [7, 8]. An early life antibiotic exposure is associated with subsequent worse neurocognitive outcomes [9, 10]. By contrast, the administration of probiotic strains yielded controversial results in terms of cognitive changes. For example, probiotic ingestion has been reported to improve sustained attention and working memory in elderly participants [11], while an early study reported potential cognitive impairments of probiotic consumption [12]. For the specific taxa, a multi-omics integration analysis revealed that three genera, *Odoribacter*, *Butyricimonas*, and *Bacteroides*, exhibited a positive association with improved cognitive performance [13]. Additionally, the abundance of *Odoribacter* was linked to several important features of brain structure and volumes [13]. Furthermore, a recent metagenomic association analysis found that bacteria with the ability to produce short-chain fatty acids (SCFAs), including *Bacteroides massiliensis*, and *Fusicatenibacter saccharivorans* were found to be positively correlated with improved cognitive performance [14]. Despite growing evidence linking gut microbiome composition and cognitive ability, the causal role is still scarce. Moreover, current conclusions are mainly based on conventional observational studies, which can be impacted by a variety of confounding factors, such as diet. It is critical to explore the potential causal role between gut microbiome composition and intelligence.

Randomized controlled trials (RCTs) of gut microbiota have the potential to establish causal relationships. Nevertheless, most RCTs are expensive and time-consuming, and more importantly, gut microbiome composition cannot be randomly allocated in practice. Alternatively, Mendelian randomization (MR) employs genetic variants as instrumental variables (IVs) to investigate the causal associations between modifiable exposures and outcomes [15]. Genetic variants are distributed randomly during meiosis and fertilization, making them largely independent of self-selected behaviors, thereby circumventing bias from confounding factors and reverse causality. Large-scale genome-wide association studies (GWASs) on the gut microbiome [16] and intelligence [17] provide the opportunity for MR analysis with significantly improved statistical power.

In the present study, we performed a bidirectional two-sample MR analysis to investigate the causal relationships between 211 gut microbiota composition (consisting of 131 genera, 35 families, 20 orders, 16 classes, and 9 phyla) and human intelligence. We eventually identified two putative causal associations. Considering the close relationship between brain volume and human intelligence [18, 19], we further conducted a two-step MR analysis to explore whether the effect of the identified taxa on intelligence was mediated by regulating the brain volume. Our findings may provide insight into the early-stage intervention of cognitive ability at the gut microbiome level.

## Methods

### Study overview

An overview of the study is shown in Figure 1. We first used single-nucleotide polymorphisms (SNPs) derived from the summary-level data as genetic instruments for the risk factor. In this study, each bacterial taxon was regarded as an independent exposure. We then performed a two-sample bidirectional MR to assess the causal effect of each gut microbiome composition on intelligence, and vice versa. At last, we utilized a two-step MR analysis to assess whether brain volume plays a causal role in mediating the pathway linking the identified gut microbiome composition and intelligence.

**Figure 1.**
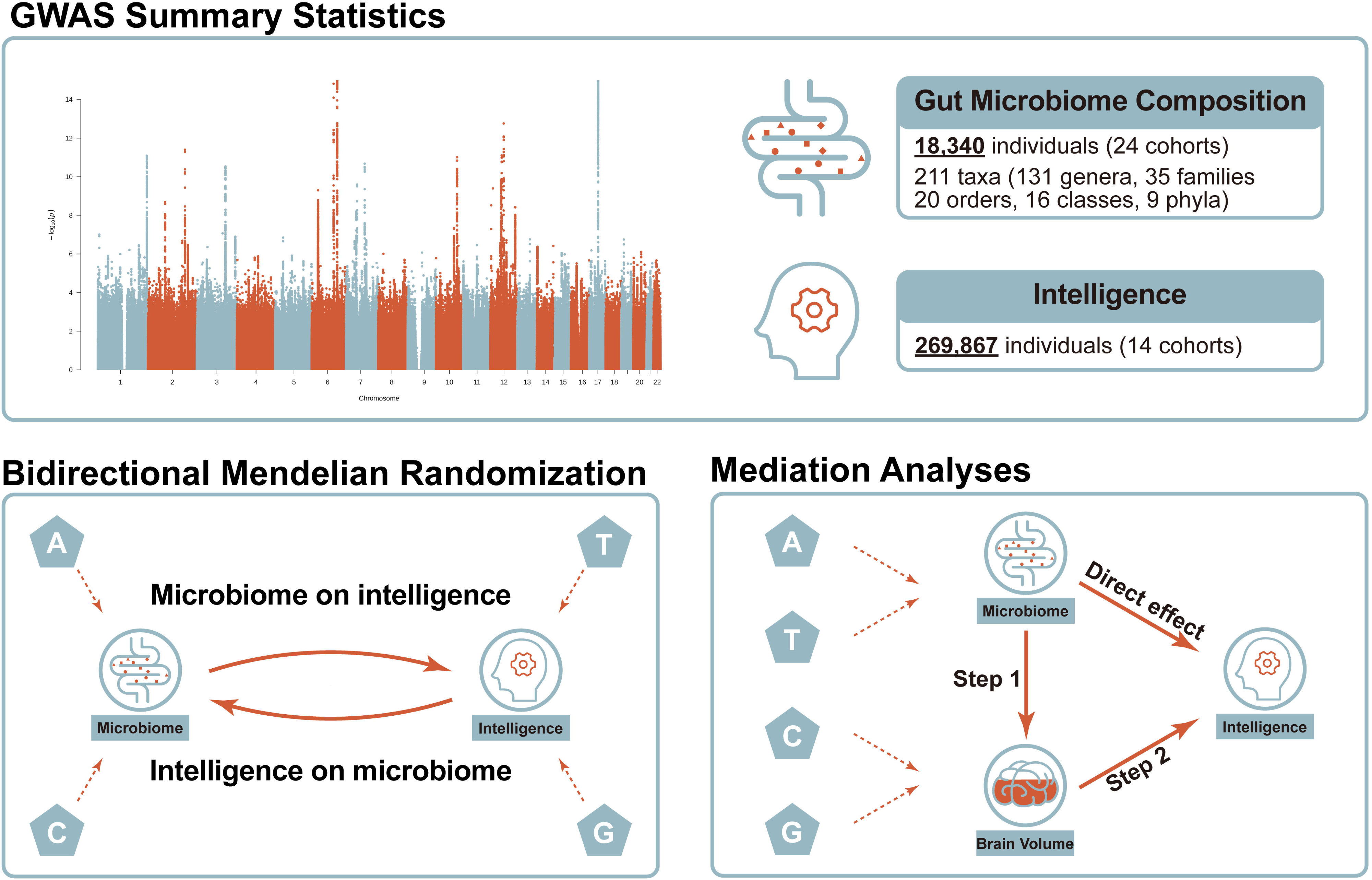
Study workflow. GWAS indicates genome-wide association study.

### Data sources

Summary information on the data sources and sample sizes used in this study can be found in Supplementary Table 1.

#### Gut microbiome composition

The genetic information for gut microbiome composition was obtained through the largest GWAS meta-analysis to date conducted by the MiBioGen consortium [16]. The study involved the coordination of 16S rRNA gene sequencing and genetic profiling of 18,340 individuals from 24 cohorts, most of whom were of European ancestry [16]. Only taxa that met the criteria of an effective sample size of at least 3,000 individuals and presence in at least three cohorts were included in the original paper. A total of 211 taxa at six levels (131 genera, 35 families, 20 orders, 16 classes, and 9 phyla) were ultimately analyzed.

#### Intelligence

The genetic associations for intelligence were derived from the largest meta-analysis of GWAS to date, which encompassed 269,867 individuals of European ancestry across 14 independent cohorts [17]. Intelligence was measured through various neurocognitive assessments, such as verbal and mathematical fluid intelligence tested in UK Biobank (UKB) [20]. But different measures were operationalized to index a common latent factor, labeled general intelligence or Spearman’s *g* [21], also known as the positive manifold of cognitive ability or intelligence. Association analysis was conducted using a linear model for all cohorts except the HiQ/HRS. These cohorts utilized a multidimensional set of cognitive performance tests to generate normally distributed scores such as a single sum, mean, or factor scores, which were subsequently employed as the phenotype. The HiQ/HRS cohort used an extreme sampling design to compare individuals with high intelligence (top 0.03% of the IQ distribution) with unascertained population controls, and a logistic regression was applied [22].

#### Brain volume

The genetic variants associated with brain volume were derived from meta-analysis results of brain volume in the UKB, as well as two additional GWAS on intracranial volume and head circumference, both considered proxy measures for brain volume [18]. A total of 17,062 participants were included in the GWAS analyses in UKB. Brain volume was estimated using structural (T_1_-weighted) magnetic resonance imaging (MRI) by combining total gray and white matter volume with ventricular cerebrospinal fluid volume. The GWAS results of intracranial volume conducted by the ENIGMA consortium included 11,373 participants [23] and head circumference included a total of 18,881 participants [24], resulting in a combined sample size of 47,316.

### Genetic instruments selection

The genetic instruments employed must fulfill three assumptions [25]: (1) the genetic variants should be strongly associated with the exposure, (2) the genetic variants should not be associated with any potential confounding factors, and (3) the genetic variants should not affect the outcome independently of exposure.

#### Selecting genetic instruments

We used the clump function within PLINK software [26] to identify independent SNPs for each exposure, using the 1000 Genomes European data as the reference. A strict cut-off of *r^2^* < 0.001, a window of 10,000 kb, along with a *P* < 5×10^-8^ were used for clumping. It is worth noting that we used a relaxed *P* value threshold of 1×10^-5^ for gut microbiome composition, similar to previous MR studies [16, 27, 28], since SNPs below this threshold were found to have the largest explained variance on microbial features [29]. To ensure consistency, we harmonized the effects of SNPs on both exposure and outcome by aligning the beta values to the same alleles. Where shared SNPs between exposure and outcome were not available, we replaced them with proxy SNPs (*r^2^* > 0.8) that were significantly associated with the exposure.

#### Removing confounders

To avoid potential confounding, we removed SNPs significantly associated with plausible confounders in the PhenoScanner database [30, 31] in European participants. Four potential confounders were taken into account, including diet, socioeconomic status, drinking, and smoking behavior. These traits have been reported to affect both gut microbiome composition [32, 33], and intelligence or cognitive ability [34–37]. In addition, SNPs associated with the outcome (*P* < 1×10^-5^) were also excluded to satisfy the third assumption.

#### Quality control of genetic instruments

We excluded palindromic SNPs with intermediate allele frequencies (>0.42), which would introduce potential strand-flipping issues. To enhance the accuracy and robustness of the remaining genetic instruments, we also removed outlier pleiotropic SNPs detected by RadialMR [38]. RadialMR identified outlier pleiotropic SNPs utilizing a heterogeneity test (modified *Q* statistics) with a nominal significance level of 0.05. *F* statistics [39] were calculated to estimate the strength of genetic instruments, and only SNPs with *F* statistics > 10 were included in the MR analysis. We only kept the results with at least three SNPs after removing confounders and quality control.

### Two-sample Mendelian randomization

Bidirectional causal relationships were performed to test if gut microbiome composition causally affects intelligence and if intelligence can causally affect gut microbiome composition. We utilized the inverse-variance weighted (IVW) method based on a multiplicative random-effects model [40] as the primary causal inference. This approach combined Wald ratio estimates from individual SNPs into a single causal estimate for each risk factor [41]. Specifically, each estimate was calculated by dividing the SNP-outcome association by the SNP-exposure association [41]. Since the IVW estimates can be biased if pleiotropic instrumental variables are introduced [42], we estimated the causality using additional four methods in parallel to enhance the reliability of our results. Briefly, robust adjusted profile score (MR-RAPS) considers systematic and idiosyncratic pleiotropy, enabling robust inference with many weak instruments [43]. The weighted-median approach can tolerate up to 50% of variants violating MR assumptions when horizontal pleiotropy is present [44]. The weighted-mode method can provide unbiased estimates if SNPs contributing to the largest cluster are valid [45]. Lastly, the MR-Egger method allows instruments to have non-zero pleiotropy and provides a way to test and estimate the pleiotropy effect in addition to causal estimates [46]. The MR estimates are expressed as β values that indicate the change in outcome units for each unit change in exposure. The *P* values from the IVW MR test were adjusted using Benjamini–Hochberg FDR correction for multiple testing to the results; for the resulting *q* value the threshold was set to 0.05. Since the large number and hierarchical structure of taxa used in our study, the multiple comparison adjustments may be excessive. We also reported nominally significant results (*P* < 0.05) in the Supplementary materials.

### Sensitivity analysis

We conducted a series of sensitivity analyses to address the potential issue of pleiotropy in the causal estimates. First, we used MR-Egger regression to assess the presence of horizontal pleiotropy based on its intercept term; deviation from zero (*P* < 0.05) was considered evidence for directional pleiotropic bias [46]. Additionally, we utilized MR-PRESSO [47] to detect the presence of pleiotropy (*P* < 0.05). MR-PRESSO compares the observed distance of all the variants to the regression line with the expected distance under the null hypothesis of no horizontal pleiotropy. Third, we assessed heterogeneity using Cochran’s Q statistic [48], which is produced by different genetic variants in the fixed-effect variance weighted analysis; a *P* value of less than 0.05 indicated the presence of pleiotropy. At last, we conducted a leave-one-out analysis to determine whether the causal association was driven by an individual variant.

### Mediation analysis

A two-step MR analysis was performed to evaluate whether the effect of identified taxa on intelligence was mediated by regulating the brain volume. In the first step, we estimated the causal effect of specific gut microbiome composition on brain volume. In the second step, we assessed the causal effect of brain volume on intelligence. The indirect effect of identified gut microbiome composition on intelligence through brain volume was evaluated using the product of coefficients method [49]. To determine the proportion of the effect of the contribution of identified taxa on intelligence that was mediated by regulating brain volume, we divided the indirect effect by the total effect. Standard errors for the indirect effect were obtained using the delta method [50].

## Results

### Causal effects of gut microbiota composition on intelligence

We conducted a two-sample MR analysis to investigate the impact of gut microbiome abundance on intelligence. The IVW analyses revealed that the genetic liability for two specific taxa, namely the genus *Oxalobacter* and genus *Fusicatenibacter*, achieved statistical significance after FDR correction. The abundance of genus *Oxalobacter* was negatively associated with intelligence (IVW beta, –0.032; 95% confidence interval (CI), –0.049 to –0.015; *P* = 1.88×10^-4^) (Table 1). The estimates were similar in size in MR RAPS (beta, –0.032; 95% CI, –0.051 to –0.014; *P* = 6.85×10^-4^). We also found causal evidence that the abundance of genus *Fusicatenibacter* was positively associated with intelligence (IVW beta, 0.051; 95% CI, 0.023 to 0.079; *P* = 3.03×10^-4^) (Table 1). The results were further supported by MR RAPS (beta, 0.052; 95% CI, 0.021 to 0.083; *P* = 1.01×10^-3^), and weighted median (beta, 0.048; 95% CI, 0.010 to 0.087; *P* = 0.014). The CIs of the weighted mode and MR Egger methods were wider compared to other methods, which could be attributed to their lower statistical power when compared to IVW [45]. Scatter plots of SNP effects on these two taxa versus intelligence are presented in Figure 2, with colored lines representing the slopes of different MR analyses. Forest plots of individual and combined SNP MR-estimated effect sizes are also presented. Moreover, we reported additional 27 taxa causally associated with intelligence at a nominally significant level (*P* < 0.05) (Supplementary Table 2).

**Figure 2.**
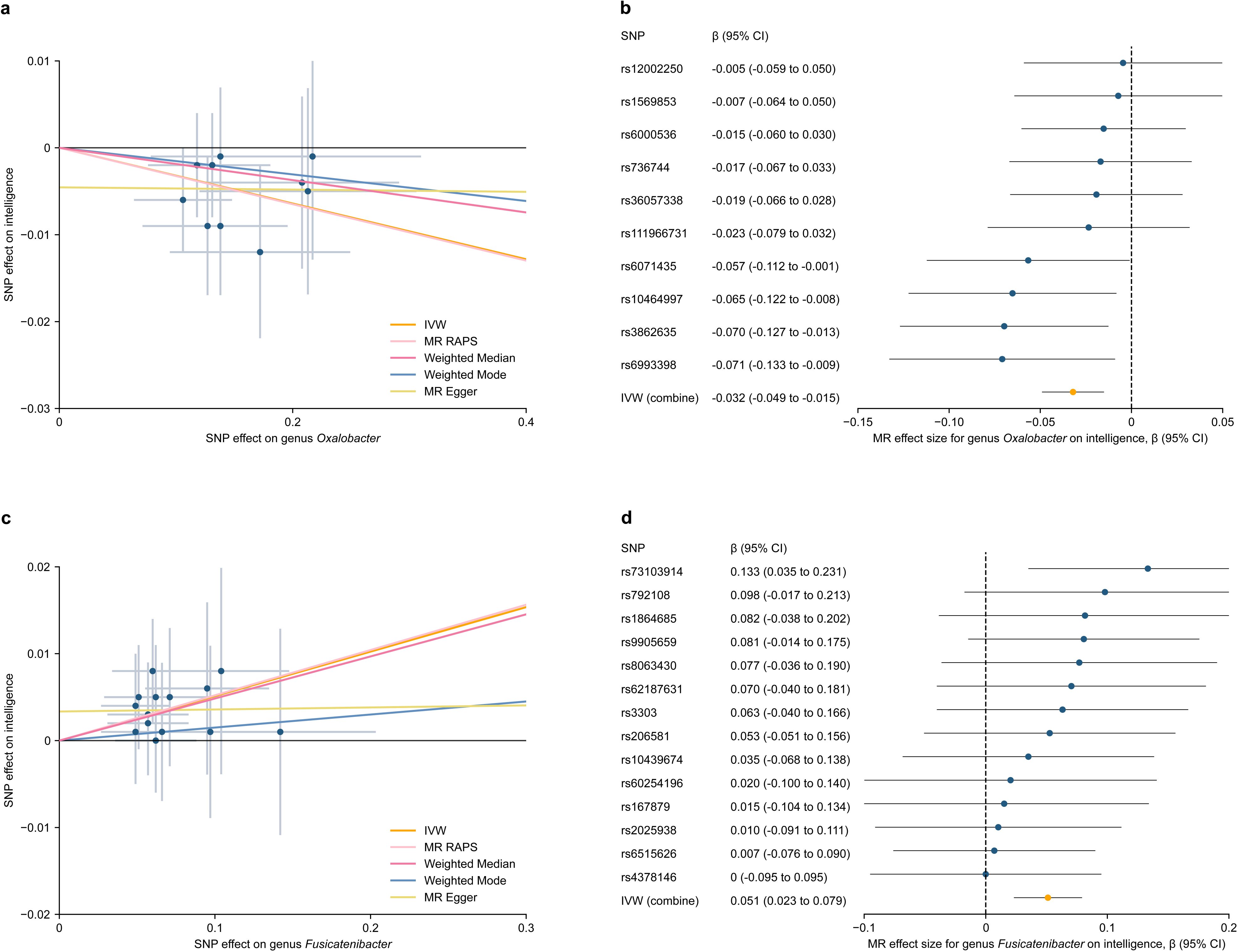
Mendelian randomization (MR) plots for the relationship of gut microbiome composition with intelligence. **a.** Scatterplot of single-nucleotide polymorphism (SNP) effects on genus *Oxalobacter* vs intelligence, with the slope of each line corresponding to estimated MR effect per method. Data are expressed as raw β values with 95% CI. IVW indicates the inverse-variance weighted method; RAPS indicates the robust adjusted profile score. **b.** Forest plot of individual and combined SNP MR-estimated effects sizes for genus *Oxalobacter* on intelligence. **c.** Scatterplot of SNP effects on genus *Fusicatenibacter* vs intelligence. **d.** Forest plot of individual and combined SNP MR-estimated effects sizes for genus *Fusicatenibacter* on intelligence.

**Table 1.**
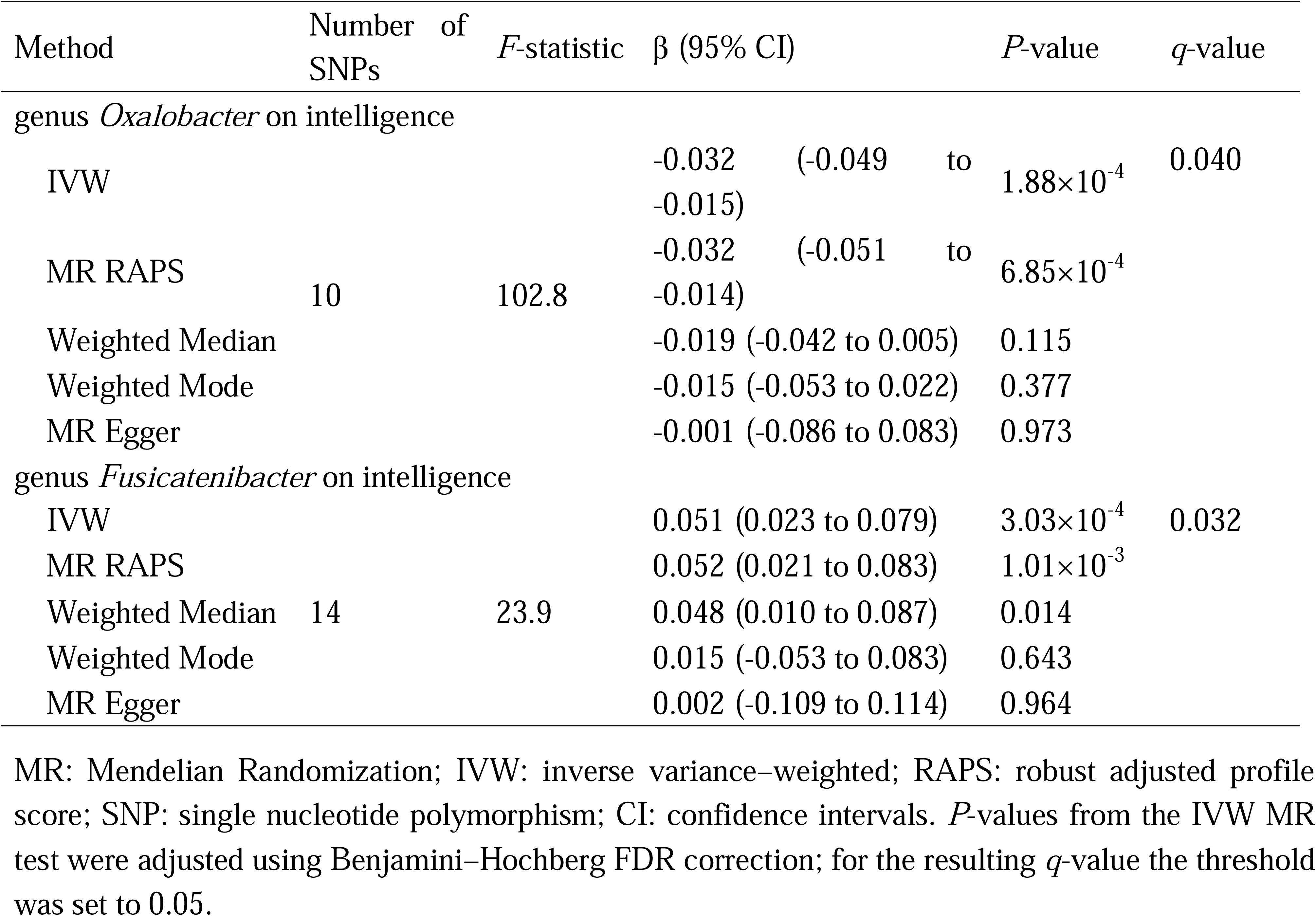
Significant MR results for the relationship between gut microbiota and intelligence.

All SNPs selected for inclusion and exclusion for the two identified taxa are presented in Supplementary Tables 3-6 for replication. After selection, 10 and 14 SNPs significantly associated with the genus *Oxalobacter* and genus *Fusicatenibacter* were used in the MR analyses. The *F* statistics for the genetic instruments indicated the absence of weak instrument bias (Table 1, Supplementary Tables 3 and 5). Sensitivity analyses did not address any pleiotropy in the causal estimates (Supplementary Table 7). Specifically, the MR-Egger intercept analysis did not reveal evidence of directional pleiotropy for the genus *Oxalobacter* (intercept, –0.005; 95%CI, –0.015 to 0.006; *P* = 0.414), and genus *Fusicatenibacter* (intercept, 0.003; 95%CI, –0.003 to 0.010; *P* = 0.341). MR-PRESSO did not detect any potential outliers for the genus *Oxalobacter* (*P* = 0.537) and genus *Fusicatenibacter* (*P* = 0.877). Furthermore, Cochran’s Q statistic indicated a lack of evidence for pleiotropy across instrument effects for the genus *Oxalobacter* (Q, 8.25; *P* = 0.509) and genus *Fusicatenibacter* (Q, 7.86; *P* = 0.853). At last, analyses leaving out each SNP found that none of the SNPs were responsible for driving the MR results (Supplementary Figure 1).

### Causal effects of intelligence on gut microbiota composition

With genetic liability for intelligence as exposure, we performed MR analyses to explore the causal effect of intelligence on the abundance of the gut microbiome. The SNPs that were included and excluded for intelligence are presented in Supplementary Tables 8-9 to replicate our findings. We found no evidence of causal relationships for intelligence on the genus *Oxalobacter* (IVW beta, 0.013; 95% CI, –0.170 to 0.196; *P* = 0.889) and genus *Fusicatenibacter* (IVW beta, 0.031; 95% CI, –0.066 to 0.127; *P* = 0.536) (Table 2, Supplementary Figure 2). Similar effect patterns were observed across the MR-RAPS, weighted median, weighted mode, and MR-Egger methods (Table 2). Additionally, the *F* statistics of the genetic instruments suggested a lack of weak instrument bias (Table 2, Supplementary Table 8). Sensitivity analyses did not address potential pleiotropy in the causal estimates (Supplementary Table 7). Analyses leaving out each SNP found that none of the SNPs were responsible for driving the MR results (Supplementary Figure 3). Although no causal evidence after multiple testing corrections, we found that the genetic liability for intelligence had a causal contribution to the abundance of 15 gut microbiomes at a nominally significant level (*P* < 0.05) (Supplementary Table 10).

**Table 2.**
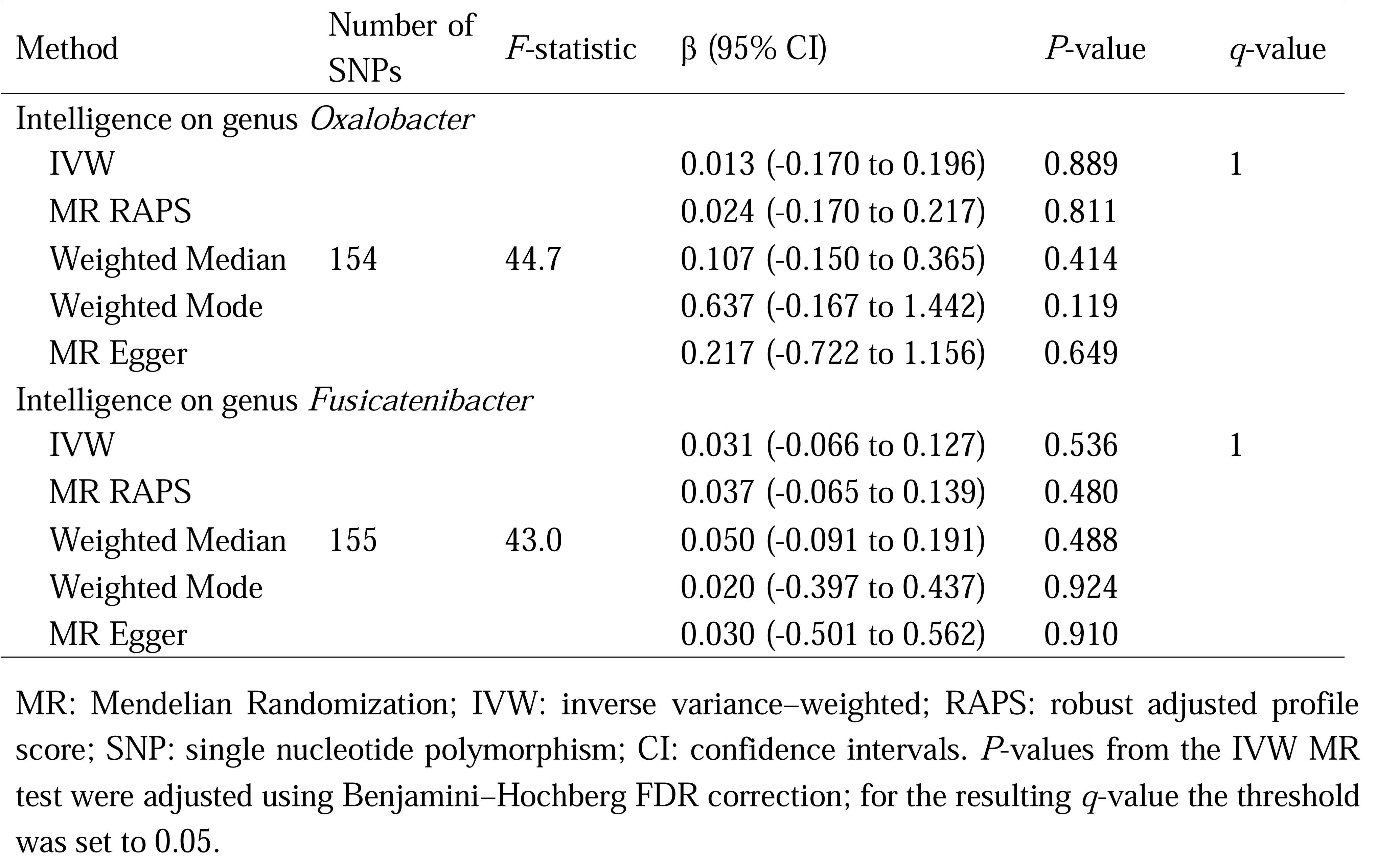
Bidirectional MR results for the relationship between intelligence and gut microbiota.

| Method | Number of SNPs | *F*-statistic | $\beta$ (95% CI) | *P*-value | *q*-value |
| --- | --- | --- | --- | --- | --- |
| Intelligence on genus *Oxalobacter* |  |  |  |  |  |
| IVW |  |  | 0.013 (-0.170 to 0.196) | 0.889 | 1 |
| MR RAPS |  |  | 0.024 (-0.170 to 0.217) | 0.811 |  |
| Weighted Median | 154 | 44.7 | 0.107 (-0.150 to 0.365) | 0.414 |  |
| Weighted Mode |  |  | 0.637 (-0.167 to 1.442) | 0.119 |  |
| MR Egger |  |  | 0.217 (-0.722 to 1.156) | 0.649 |  |
| Intelligence on genus *Fusicatenibacter* |  |  |  |  |  |
| IVW |  |  | 0.031 (-0.066 to 0.127) | 0.536 | 1 |
| MR RAPS |  |  | 0.037 (-0.065 to 0.139) | 0.480 |  |
| Weighted Median | 155 | 43.0 | 0.050 (-0.091 to 0.191) | 0.488 |  |
| Weighted Mode |  |  | 0.020 (-0.397 to 0.437) | 0.924 |  |
| MR Egger |  |  | 0.030 (-0.501 to 0.562) | 0.910 |  |

MR: Mendelian Randomization; IVW: inverse variance-weighted; RAPS: robust adjusted profile score; SNP: single nucleotide polymorphism; CI: confidence intervals. *P*-values from the IVW MR test were adjusted using Benjamini–Hochberg FDR correction; for the resulting *q*-value the threshold was set to 0.05.

### Mediation analysis

Considering the close relationship between brain volume on human intelligence [18, 19], we performed a two-step MR analysis to investigate whether the effect of identified taxa on intelligence was mediated by regulating the brain volume.

We first estimated the causal effect of identified taxa on brain volume using genetic instruments specific to the genus *Oxalobacter* and genus *Fusicatenibacter*. Summary information on brain volume for the SNPs associated with these two taxa is listed in Supplementary Tables 11-12. We identified that increased genus *Fusicatenibacter* was associated with increased brain volume (IVW beta, 0.086; 95% CI, 0.019 to 0.153; *P* = 0.012) (Figure 3a, Supplementary Figure 4). We found no evidence of causal relationships for genus *Oxalobacter* on the brain volume (IVW beta, 0.006; 95% CI, –0.033 to 0.046; *P* = 0.745). Sensitivity analyses did not address any pleiotropy in the causal estimates (Supplementary Table 13). Analyses leaving out each SNP found that none of the SNPs were responsible for driving the MR results (Supplementary Figure 5).

**Figure 3.**
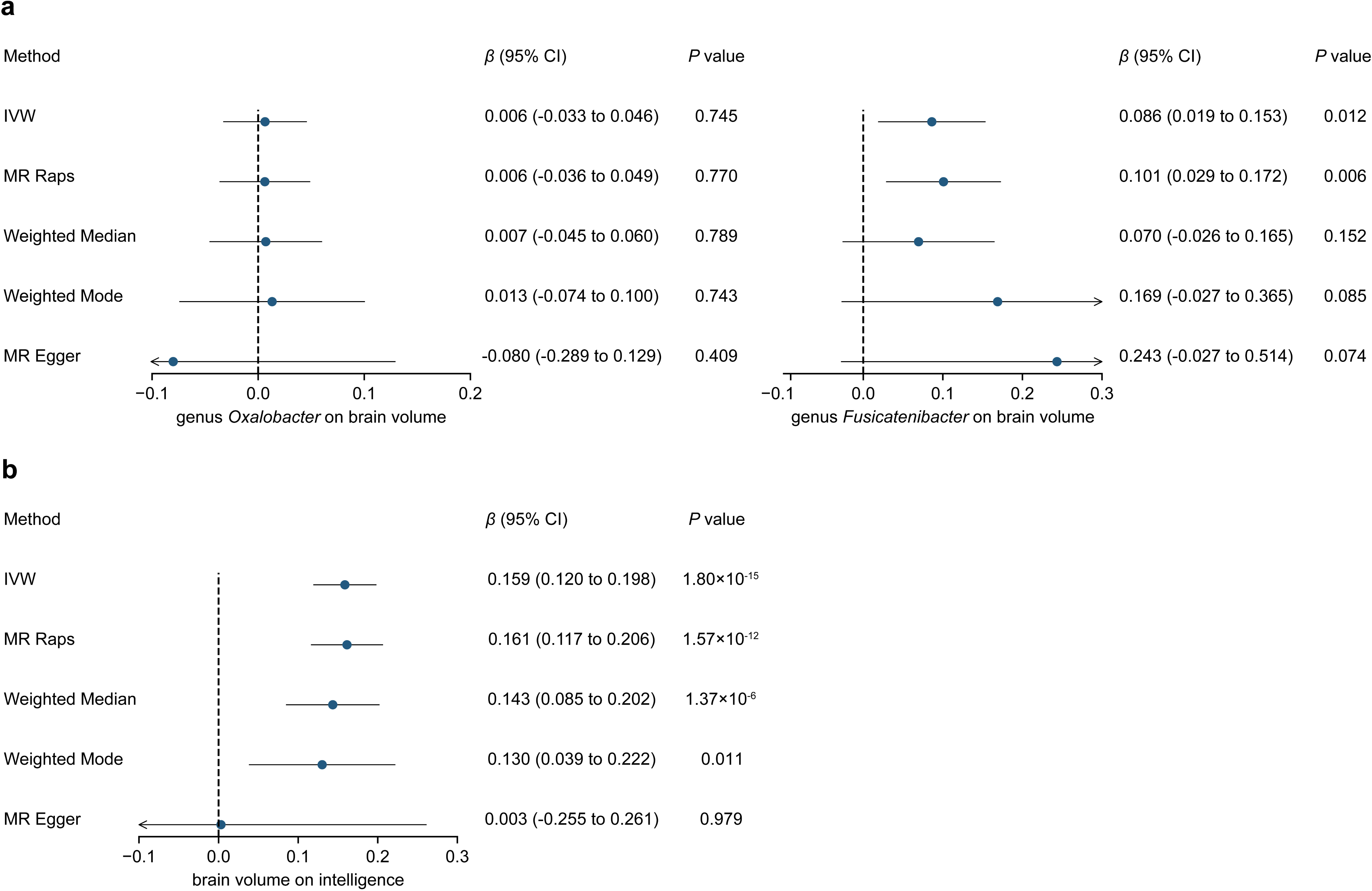
Mediation analysis of gut microbiome composition on intelligence via brain volume. **a.** Summary MR estimates derived from the inverse-variance weighted (IVW), weighted median, weighted mode, robust adjusted profile score (MR RAPS), and MR-Egger methods for genus *Oxalobacter* on brain volume (left) and genus *Fusicatenibacter* on brain volume (right). **b.** Summary MR estimates for brain volume on intelligence.

We then assessed the causal effect of brain volume on intelligence using genetic instruments that were associated with brain volume (Supplementary Tables 14-15). We found extremely causal evidence for the effects of brain volume on intelligence (IVW beta, 0.159; 95% CI, 0.120 to 0.198; *P*

= 1.80×10^-15^) (Figure 3b, Supplementary Figure 6), similar to the results reported in a previous paper [18]. Sensitivity analyses did not address any pleiotropy in the causal estimates (Supplementary Table 13). Analyses leaving out each SNP found that none of the SNPs were responsible for driving the MR results (Supplementary Figure 7).

Finally, we revealed that the genus *Fusicatenibacter* indirectly affects intelligence by regulating brain volume. Specifically, the mediation effect was estimated to be 0.014 (95%CI, 0.003 to 0.025; *P* = 0.016) with a mediation proportion of 26.7% (95% CI, 4.9% to 48.5%) (Table 3).

**Table 3.**
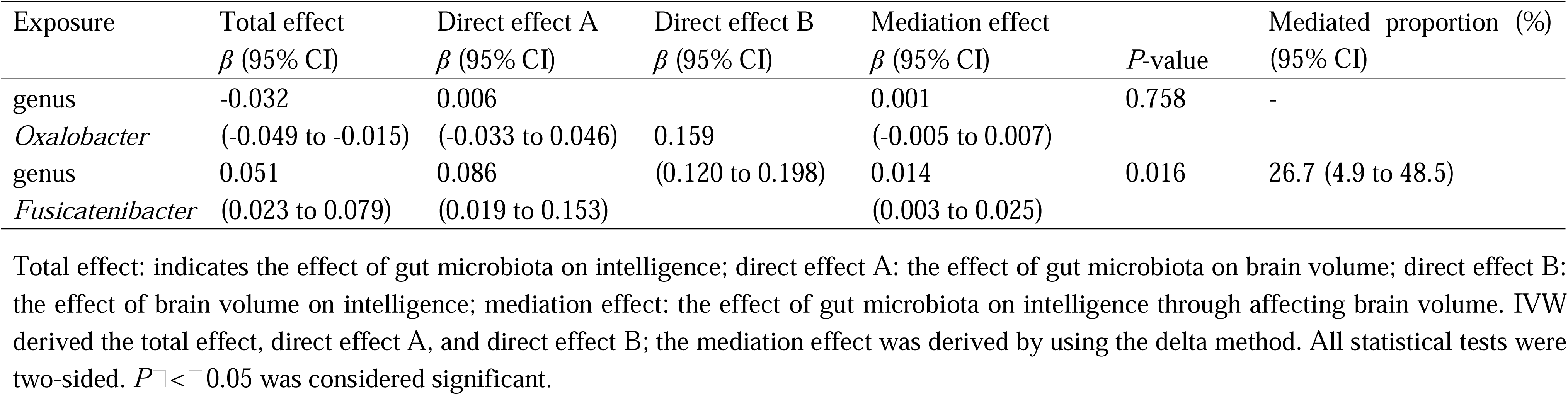
The mediation effect of gut microbiota on intelligence via affecting brain volume.

## Discussion

The role of our microbiome play in health and disease is one of the biggest scientific challenges [51]. In this study, by utilizing summary statistics obtained from the largest GWAS meta-analysis of gut microbiota and intelligence, we conducted a bidirectional two-sample MR analysis to disentangle the causal relationship. We observed causal evidence indicating a risk effect of the genus *Oxalobacter* and a protective effect of the genus *Fusicatenibacter* on intelligence. As for the other direction, we found no evidence of causal relationships of intelligence with the abundance of the gut microbiome. More interestingly, we conducted a mediation analysis and showed that the effect of genus *Fusicatenibacter* on intelligence was partially mediated by regulating brain volume. The findings may have implications for public health interventions that seek to enhance individual intelligence.

The gut microbiota potentially impacts the development and function of the immune, metabolic, and nervous systems through bidirectional communication along the gut-brain axis [52], which is believed to be involved in the intelligence/cognitive ability of the host [8]. For instance, compared to normal mice, germ-free mice showed impairments in tests of memory and reductions in hippocampal brain-derived neurotrophic factor (BDNF) [53], which is a neurotrophin crucial for neuronal development and survival, synaptic plasticity, and cognitive function. By contrast, the administration of probiotic strains can promote memory behavior [54] through their production of lactate and the promotion of gamma-aminobutyric acid (GABA) accumulation in the hippocampus [55], providing solid evidence for the role of the microbiome in intelligence. The association between the gut microbiome and cognitive ability in animals also occurs in humans as mentioned in the introduction. However, current conclusions predominantly rely on disparities in gut microbiome composition and the results of trials that involved the transplant of gut microbiota into gnotobiotic mice [56–58], which can be impacted by various confounding factors. As the gut microbiome is considered to be highly dynamic, causal association has been an unresolved issue in the field.

To our knowledge, we report the first MR analysis to investigate the potential causal relationship between gut microbiota and intelligence. We found the genetic liability for two taxa, genus *Oxalobacter* and genus *Fusicatenibacter*, reached a statistical significance after FDR correction. *Oxalobacter* is one of the key taxa involved in the gut microbiome diversity of individuals [59], and previous studies showed that *Oxalobacter formigenes* plays an important role in oxalate absorption and secretion pathways in the gut [60]. Observational studies found controversial associations between the genus *Oxalobacter* and cognitive ability. Some studies reported a negative association between the abundance of the genus *Oxalobacter* and the cognitive ability estimated by the Mini-Mental State Examination (MMSE) score [61]. On the contrary, another study reported the reduced abundance of the genus *Oxalobacter* was associated with mild cognitive impairment in older adults [62]. Our MR analysis observed causal evidence indicating the risk effect of the genus *Oxalobacter* on intelligence. We also provided the protective effect of the genus *Fusicatenibacter* on intelligence. A recent cross-sectional analysis focusing on species-level features associated with cognition found that certain bacteria capable of producing short-chain fatty acids, such as *Fusicatenibacter saccharivorans*, were positively associated with better cognitive performance [14]. Specifically, a higher abundance of *Fusicatenibacter saccharivorans* was linked to better scores on both the MMSE and the Montreal Cognitive Assessment (MoCA) [14].

Another interesting conclusion arising from the current study is that the protective effect of genus *Fusicatenibacter* abundance on intelligence was partially mediated by increasing brain volume. In the first step, we identified an increased abundance of the genus *Fusicatenibacter* associated with increased intelligence. Differences in gut microbial composition have been reported associated with brain structure [63–66]. Specifically, multimodal neuroimaging fusion biomarkers have been reported to mediate the association between gut microbiota and cognition [67]. Yet little is known about the effect of genus *Fusicatenibacter* on brain volume. The second step provided evidence of genetically determined higher brain volume was associated with higher intelligence. Jansen and colleagues [18] found causal evidence of genetically predicted brain volume on intelligence (beta = 0.154, *P* = 1.88× 10^-23^) using the generalized summary-data-based MR (GSMR) package. This result supports our second-step estimate in terms of both direction and magnitude.

Admittedly, several limitations should be acknowledged when interpreting the results of this study. First, although we utilized the largest GWAS meta-analysis for gut microbiome composition to date, the number of subjects in gut microbiome composition GWAS is relatively small, and genetic factors can only explain a small proportion of variance in gut microbiome features; thus, the power to detect the causal relationship was limited. Second, we used a relaxed *P* value threshold to select genetic instruments due to the limited variant number associated with gut microbiome composition, in line with previous microbiota MR studies [16, 27, 28], which may introduce weak instrument bias. Nevertheless, we tested the instrument strength and found that all instruments enjoyed *F* statistics exceeding 10, a conventional cutoff for strong instruments [68]. Third, due to the lowest taxonomic level being genus in the exposure dataset, we were unable to explore the causal association between gut microbiota and intelligence at the species level. Finally, although the majority of participants in the GWAS summary data were European, a small number of the gut microbiota data from other races may result in weak instrument variables and potential bias estimates [69]. Moreover, it might difficult to extrapolate our results to different ethnic populations, and significant replication when diverse samples become available is essential.

## Conclusion

In summary, we used the largest GWAS meta-analysis of gut microbiota and intelligence to disentangle the causal association. We found robust genetic evidence of gut microbiome features on intelligence, and the protective effect of genus *Fusicatenibacter* on intelligence was partially mediated by regulating brain volume. Our findings may potentially reshape our understanding of the microbiota-gut-brain axis and highlight the gut microbiota as a prospective target for treating and preventing cognitive impairment.

## Data availability

All GWAS summary statistics analyzed in the current study could be downloaded from the public domain. The GWAS for gut microbiome composition can be obtained through the NHGRI-EBI GWAS catalog (https://www.ebi.ac.uk/gwas), study accession nos. GCST90016908-GCST90017118. The GWAS summary statistics for intelligence and brain volume are available for download at the website of the Department of Complex Trait Genetics (https://ctg.cncr.nl/software/summary_statistics/). All data generated in the current study could be obtained from the Supplementary Information.

## Code availability

All the analyses used in this study were conducted using the R packages TwoSampleMR (version 0.4.21), RadialMR (version 0.4), and MR-PRESSO (version 1.0). The code to reproduce all results reported in the manuscript is available from the corresponding author upon reasonable request.

## Data Availability

All data produced in the present work are contained in the manuscript

