## Supplementary Table 1 for "Mendelian Randomization analyses identify causal associations of human gut microbiome composition on intelligence"

**Supplementary Table 1. GWAS summary statistics: source and description**

| **Phenotypes** | **Sample size** | **Reference (PMID)** | **Download** |
| --- | --- | --- | --- |
| Gut Microbiota | 18,340 | 33462485 | NHGRI-EBI GWAS Catalog  (nos. GCST90016908 to GCST90017118) |
| Intelligence | 269,867 | 29942086 | https://ctg.cncr.nl/documents/p1651/SavageJansen_IntMeta_sumstats.zip |
| Brain volume | 47,316 | 33154357 | https://ctg.cncr.nl/documents/p1651/meta_analysis_BV_Jansenetal_2020.sumstats.txt.gz |

**Supplementary Table 2. Significant MR results (*P* < 0.05) for the relationship between gut microbiota and intelligence**

| **Method** | **Number of SNPs** | ***F*-statistic** | **β (95% CI)** | ***P*-value** | ***q*-value** |
| --- | --- | --- | --- | --- | --- |
| genus *Oxalobacter* on intelligence | | | | | |
| IVW | 10 | 102.8 | -0.032 (-0.049 to -0.015) | 1.88×10^-4^ | 0.040 |
| MR RAPS |  |  | -0.032 (-0.051 to -0.014) | 6.85×10^-4^ |  |
| Weighted Median |  |  | -0.019 (-0.042 to 0.005) | 0.115 |  |
| Weighted Mode |  |  | -0.015 (-0.053 to 0.022) | 0.377 |  |
| MR Egger |  |  | -0.001 (-0.086 to 0.083) | 0.973 |  |
| genus *Fusicatenibacter* on intelligence | | | | | |
| IVW | 14 | 23.9 | 0.051 (0.023 to 0.079) | 3.03×10^-4^ | 0.032 |
| MR RAPS |  |  | 0.052 (0.021 to 0.083) | 0.001 |  |
| Weighted Median |  |  | 0.048 (0.010 to 0.087) | 0.014 |  |
| Weighted Mode |  |  | 0.015 (-0.053 to 0.083) | 0.643 |  |
| MR Egger |  |  | 0.002 (-0.109 to 0.114) | 0.872 |  |
| genus *Ruminococcaceae UCG003* on intelligence | | | | | |
| IVW | 12 | 30 | 0.046 (0.019 to 0.072) | 7.44×10^-4^ | 0.052 |
| MR RAPS |  |  | 0.048 (0.018 to 0.077) | 0.001 |  |
| Weighted Median |  |  | 0.047 (0.009 to 0.084) | 0.014 |  |
| Weighted Mode |  |  | 0.073 (0.003 to 0.142) | 0.042 |  |
| MR Egger |  |  | 0.014 (-0.084 to 0.113) | 0.751 |  |
| genus *Candidatus Soleaferrea* on intelligence | | | | | |
| IVW | 10 | 64.6 | -0.031 (-0.051 to -0.011) | 0.003 | 0.134 |
| MR RAPS |  |  | -0.031 (-0.054 to -0.009) | 0.006 |  |
| Weighted Median |  |  | -0.033 (-0.059 to -0.007) | 0.012 |  |
| Weighted Mode |  |  | -0.034 (-0.077 to 0.010) | 0.113 |  |
| MR Egger |  |  | -0.019 (-0.097 to 0.060) | 0.603 |  |
| genus *Roseburia* on intelligence | | | | | |
| IVW | 15 | 26.5 | -0.040 (-0.067 to -0.014) | 0.003 | 0.132 |
| MR RAPS |  |  | -0.043 (-0.072 to -0.013) | 0.004 |  |
| Weighted Median |  |  | -0.043 (-0.080 to -0.006) | 0.023 |  |
| Weighted Mode |  |  | -0.051 (-0.120 to 0.017) | 0.129 |  |
| MR Egger |  |  | -0.036 (-0.115 to 0.043) | 0.342 |  |
| genus *Dialister* on intelligence | | | | | |
| IVW | 10 | 32.5 | 0.042 (0.013 to 0.070) | 0.004 | 0.139 |
| MR RAPS |  |  | 0.044 (0.012 to 0.075) | 0.007 |  |
| Weighted Median |  |  | 0.032 (-0.006 to 0.070) | 0.097 |  |
| Weighted Mode |  |  | 0.031 (-0.040 to 0.101) | 0.356 |  |
| MR Egger |  |  | 0.096 (-0.041 to 0.233) | 0.144 |  |
| genus *Phascolarctobacterium* on intelligence | | | | | |
| IVW | 11 | 37.7 | -0.037 (-0.062 to -0.012) | 0.004 | 0.129 |
| MR RAPS |  |  | -0.037 (-0.065 to -0.010) | 0.008 |  |
| Weighted Median |  |  | -0.026 (-0.061 to 0.010) | 0.154 |  |
| Weighted Mode |  |  | -0.017 (-0.080 to 0.046) | 0.561 |  |
| MR Egger |  |  | 0.004 (-0.101 to 0.109) | 0.931 |  |
| genus *Holdemanella* on intelligence | | | | | |
| IVW | 10 | 53.3 | 0.035 (0.010 to 0.060) | 0.006 | 0.168 |
| MR RAPS |  |  | 0.038 (0.011 to 0.065) | 0.007 |  |
| Weighted Median |  |  | 0.026 (-0.006 to 0.058) | 0.112 |  |
| Weighted Mode |  |  | 0.064 (3.75×10^-4^ to 0.128) | 0.049 |  |
| MR Egger |  |  | 0.219 (-0.079 to 0.516) | 0.129 |  |
| genus *Lachnospiraceae UCG001* on intelligence | | | | | |
| IVW | 12 | 39.9 | 0.032 (0.008 to 0.056) | 0.008 | 0.190 |
| MR RAPS |  |  | 0.033 (0.007 to 0.060) | 0.015 |  |
| Weighted Median |  |  | 0.028 (-0.003 to 0.059) | 0.078 |  |
| Weighted Mode |  |  | 0.016 (-0.047 to 0.078) | 0.588 |  |
| MR Egger |  |  | -0.038 (-0.183 to 0.108) | 0.577 |  |
| genus *Ruminococcaceae NK4A214 group* on intelligence | | | | | |
| IVW | 12 | 27.2 | -0.037 (-0.066 to -0.009) | 0.010 | 0.201 |
| MR RAPS |  |  | -0.040 (-0.072 to -0.009) | 0.012 |  |
| Weighted Median |  |  | -0.046 (-0.087 to -0.005) | 0.027 |  |
| Weighted Mode |  |  | -0.048 (-0.116 to 0.019) | 0.143 |  |
| MR Egger |  |  | -0.040 (-0.128 to 0.047) | 0.331 |  |
| order *Clostridiales* on intelligence | | | | | |
| IVW | 15 | 20.6 | 0.038 (0.009 to 0.067) | 0.010 | 0.186 |
| MR RAPS |  |  | 0.042 (0.010 to 0.074) | 0.011 |  |
| Weighted Median |  |  | 0.051 (0.009 to 0.092) | 0.018 |  |
| Weighted Mode |  |  | 0.077 (-0.010 to 0.164) | 0.079 |  |
| MR Egger |  |  | 0.134 (4.45×10^-4^ to 0.267) | 0.049 |  |
| genus *Catenibacterium* on intelligence | | | | | |
| IVW | 5 | 116 | -0.028 (-0.049 to -0.007) | 0.010 | 0.171 |
| MR RAPS |  |  | -0.028 (-0.051 to -0.004) | 0.021 |  |
| Weighted Median |  |  | -0.022 (-0.050 to 0.006) | 0.125 |  |
| Weighted Mode |  |  | -0.017 (-0.072 to 0.037) | 0.427 |  |
| MR Egger |  |  | -0.039 (-0.375 to 0.296) | 0.735 |  |
| genus *Lachnospiraceae UCG010* on intelligence | | | | | |
| IVW | 9 | 36.3 | 0.037 (0.008 to 0.066) | 0.011 | 0.184 |
| MR RAPS |  |  | 0.039 (0.008 to 0.071) | 0.014 |  |
| Weighted Median |  |  | 0.043 (0.006 to 0.081) | 0.025 |  |
| Weighted Mode |  |  | 0.067 (-0.010 to 0.145) | 0.080 |  |
| MR Egger |  |  | 0.113 (0.024 to 0.202) | 0.020 |  |
| genus unknown id.2071 on intelligence | | | | | |
| IVW | 14 | 32.8 | -0.030 (-0.054 to -0.005) | 0.016 | 0.247 |
| MR RAPS |  |  | -0.029 (-0.056 to -0.003) | 0.029 |  |
| Weighted Median |  |  | -0.019 (-0.055 to 0.016) | 0.289 |  |
| Weighted Mode |  |  | -0.006 (-0.067 to 0.055) | 0.842 |  |
| MR Egger |  |  | 0.077 (-0.051 to 0.204) | 0.215 |  |
| genus *Lachnospiraceae NK4A136 group* on intelligence | | | | | |
| IVW | 13 | 32.7 | -0.031 (-0.056 to -0.006) | 0.017 | 0.234 |
| MR RAPS |  |  | -0.034 (-0.062 to -0.006) | 0.018 |  |
| Weighted Median |  |  | -0.036 (-0.071 to -0.002) | 0.039 |  |
| Weighted Mode |  |  | -0.040 (-0.089 to 0.008) | 0.096 |  |
| MR Egger |  |  | -0.038 (-0.094 to 0.018) | 0.167 |  |
| genus *Collinsella* on intelligence | | | | | |
| IVW | 8 | 27.3 | -0.042 (-0.077 to -0.007) | 0.017 | 0.226 |
| MR RAPS |  |  | -0.043 (-0.081 to -0.004) | 0.031 |  |
| Weighted Median |  |  | -0.034 (-0.078 to 0.010) | 0.133 |  |
| Weighted Mode |  |  | -0.030 (-0.111 to 0.052) | 0.418 |  |
| MR Egger |  |  | -0.083 (-0.237 to 0.071) | 0.234 |  |
| genus *Eubacterium xylanophilum group* on intelligence | | | | | |
| IVW | 9 | 31.1 | 0.037 (0.006 to 0.068) | 0.019 | 0.230 |
| MR RAPS |  |  | 0.038 (0.004 to 0.072) | 0.031 |  |
| Weighted Median |  |  | 0.036 (-0.005 to 0.077) | 0.086 |  |
| Weighted Mode |  |  | 0.034 (-0.035 to 0.103) | 0.290 |  |
| MR Egger |  |  | 0.035 (-0.075 to 0.145) | 0.478 |  |
| genus *Ruminococcaceae UCG004* on intelligence | | | | | |
| IVW | 9 | 40.3 | 0.032 (0.005 to 0.058) | 0.020 | 0.235 |
| MR RAPS |  |  | 0.034 (0.004 to 0.063) | 0.024 |  |
| Weighted Median |  |  | 0.026 (-0.011 to 0.063) | 0.163 |  |
| Weighted Mode |  |  | 0.030 (-0.038 to 0.097) | 0.344 |  |
| MR Egger |  |  | -0.057 (-0.237 to 0.123) | 0.481 |  |
| family *Streptococcaceae* on intelligence | | | | | |
| IVW | 15 | 25.6 | 0.030 (0.004 to 0.056) | 0.023 | 0.253 |
| MR RAPS |  |  | 0.032 (0.003 to 0.060) | 0.029 |  |
| Weighted Median |  |  | 0.024 (-0.012 to 0.060) | 0.187 |  |
| Weighted Mode |  |  | 0.021 (-0.047 to 0.089) | 0.522 |  |
| MR Egger |  |  | -0.009 (-0.125 to 0.108) | 0.877 |  |
| genus *Anaerofilum* on intelligence | | | | | |
| IVW | 7 | 80.9 | 0.024 (0.002 to 0.045) | 0.030 | 0.313 |
| MR RAPS |  |  | 0.024 (4.99×10^-4^ to 0.048) | 0.045 |  |
| Weighted Median |  |  | 0.020 (-0.009 to 0.049) | 0.181 |  |
| Weighted Mode |  |  | 0.016 (-0.039 to 0.071) | 0.508 |  |
| MR Egger |  |  | 0.071 (-0.065 to 0.207) | 0.238 |  |
| genus *Blautia* on intelligence | | | | | |
| IVW | 10 | 25.5 | -0.035 (-0.067 to -0.003) | 0.033 | 0.326 |
| MR RAPS |  |  | -0.034 (-0.069 to 0.001) | 0.060 |  |
| Weighted Median |  |  | -0.030 (-0.074 to 0.014) | 0.179 |  |
| Weighted Mode |  |  | -0.026 (-0.100 to 0.045) | 0.428 |  |
| MR Egger |  |  | -0.016 (-0.109 to 0.078) | 0.709 |  |
| order *Lactobacillales* on intelligence | | | | | |
| IVW | 15 | 30.2 | 0.027 (0.002 to 0.052) | 0.033 | 0.313 |
| MR RAPS |  |  | 0.027 (-3.92×10^-4^ to 0.054) | 0.053 |  |
| Weighted Median |  |  | 0.022 (-0.012 to 0.055) | 0.212 |  |
| Weighted Mode |  |  | 0.019 (-0.033 to 0.070) | 0.451 |  |
| MR Egger |  |  | 0.015 (-0.047 to 0.078) | 0.605 |  |
| genus *Victivallis* on intelligence | | | | | |
| IVW | 10 | 114 | -0.016 (-0.031 to -8.86×10^-4^) | 0.038 | 0.349 |
| MR RAPS |  |  | -0.017 (-0.033 to -1.41×10^-4^) | 0.048 |  |
| Weighted Median |  |  | -0.012 (-0.033 to 0.010) | 0.284 |  |
| Weighted Mode |  |  | -0.013 (-0.053 to 0.027) | 0.485 |  |
| MR Egger |  |  | -0.045 (-0.157 to 0.068) | 0.387 |  |
| genus *Ruminococcus gauvreauii group* on intelligence | | | | | |
| IVW | 10 | 29.2 | 0.033 (0.002 to 0.064) | 0.038 | 0.337 |
| MR RAPS |  |  | 0.036 (0.002 to 0.069) | 0.038 |  |
| Weighted Median |  |  | 0.035 (-0.006 to 0.076) | 0.098 |  |
| Weighted Mode |  |  | 0.067 (-0.024 to 0.157) | 0.129 |  |
| MR Egger |  |  | -0.053 (-0.294 to 0.188) | 0.624 |  |
| genus *Eggerthella* on intelligence | | | | | |
| IVW | 7 | 73.5 | -0.023 (-0.046 to -0.001) | 0.041 | 0.343 |
| MR RAPS |  |  | -0.025 (-0.050 to -7.36×10^-5^) | 0.049 |  |
| Weighted Median |  |  | -0.030 (-0.059 to -4.47×10^-4^) | 0.047 |  |
| Weighted Mode |  |  | -0.040 (-0.095 to 0.014) | 0.120 |  |
| MR Egger |  |  | -0.039 (-0.194 to 0.115) | 0.541 |  |
| class *Mollicutes* on intelligence | | | | | |
| IVW | 8 | 35.8 | -0.033 (-0.065 to -0.001) | 0.041 | 0.332 |
| MR RAPS |  |  | -0.035 (-0.070 to 3.46×10^-4^) | 0.052 |  |
| Weighted Median |  |  | -0.026 (-0.070 to 0.018) | 0.245 |  |
| Weighted Mode |  |  | -0.019 (-0.104 to 0.066) | 0.615 |  |
| MR Egger |  |  | -0.015 (-0.236 to 0.206) | 0.873 |  |
| phylum *Tenericutes* on intelligence | | | | | |
| IVW | 8 | 35.8 | -0.033 (-0.065 to -0.001) | 0.041 | 0.319 |
| MR RAPS |  |  | -0.035 (-0.070 to 3.46×10^-4^) | 0.052 |  |
| Weighted Median |  |  | -0.026 (-0.069 to 0.017) | 0.231 |  |
| Weighted Mode |  |  | -0.019 (-0.105 to 0.067) | 0.619 |  |
| MR Egger |  |  | -0.015 (-0.236 to 0.206) | 0.873 |  |
| genus *Ruminococcus torques group* on intelligence | | | | | |
| IVW | 10 | 23.4 | 0.035 (9.41×10^-4^ to 0.068) | 0.044 | 0.331 |
| MR RAPS |  |  | 0.039 (0.002 to 0.076) | 0.038 |  |
| Weighted Median |  |  | 0.044 (-0.004 to 0.092) | 0.071 |  |
| Weighted Mode |  |  | 0.047 (-0.030 to 0.125) | 0.202 |  |
| MR Egger |  |  | 0.113 (-0.035 to 0.261) | 0.116 |  |
| genus *Romboutsia* on intelligence | | | | | |
| IVW | 10 | 39.5 | 0.027 (5.52×10^-4^ to 0.053) | 0.045 | 0.330 |
| MR RAPS |  |  | 0.029 (-4.92×10^-4^ to 0.058) | 0.054 |  |
| Weighted Median |  |  | 0.028 (-0.007 to 0.064) | 0.120 |  |
| Weighted Mode |  |  | 0.028 (-0.021 to 0.077) | 0.231 |  |
| MR Egger |  |  | 0.026 (-0.048 to 0.101) | 0.435 |  |

MR: Mendelian Randomization; IVW: inverse variance–weighted; RAPS: robust adjusted profile score; SNP: single nucleotide polymorphism; CI: confidence intervals. *P*-values from the IVW MR test were adjusted using Benjamini–Hochberg FDR correction.

**Supplementary Table 3. Summary information on genus *Oxalobacter* SNPs used as genetic instruments for the Mendelian randomization analyses**

| **SNP** | **A1** | **A2** | **EAF** | **Beta** | **SE** | **N** | ***P*-value** | **R^2^ (%)** | **F-statistics** |
| --- | --- | --- | --- | --- | --- | --- | --- | --- | --- |
| rs4428215 | G | A | 0.260 | 0.130 | 0.024 | 4655 | 7.51E-08 | 0.654 | 30.6 |
| rs736744 | C | T | 0.416 | 0.118 | 0.021 | 4655 | 2.57E-08 | 0.675 | 31.6 |
| rs6000536 | C | T | 0.211 | -0.131 | 0.025 | 4654 | 2.06E-07 | 0.571 | 26.7 |
| rs36057338 | G | T | 0.076 | 0.208 | 0.042 | 4244 | 8.80E-07 | 0.603 | 25.8 |
| rs6071435 | T | A | 0.364 | -0.106 | 0.021 | 4635 | 1.07E-06 | 0.515 | 24.0 |
| rs12002250 | A | C | 0.060 | 0.217 | 0.047 | 4297 | 1.42E-06 | 0.529 | 22.8 |
| rs1569853 | T | C | 0.138 | -0.138 | 0.030 | 4492 | 3.65E-06 | 0.454 | 20.5 |
| rs11108500 | A | G | 0.077 | -0.199 | 0.043 | 4303 | 3.74E-06 | 0.560 | 24.2 |
| rs10464997 | G | A | 0.153 | 0.138 | 0.029 | 4650 | 3.30E-06 | 0.492 | 23.0 |
| rs111966731 | T | C | 0.072 | 0.213 | 0.047 | 3931 | 7.30E-06 | 0.604 | 23.9 |
| rs6993398 | G | A | 0.153 | 0.127 | 0.028 | 4656 | 7.13E-06 | 0.420 | 19.6 |
| rs3862635 | C | T | 0.079 | -0.172 | 0.039 | 4469 | 9.19E-06 | 0.429 | 19.2 |

SNP: single nucleotide polymorphism; A1: effect allele; A2: baseline allele; EAF: effect allele frequency; SE: standard error; R^2^: Explained incremental variance in the phenotype.

**Supplementary Table 4. Summary information on intelligence for the 12 genome-wide significant SNPs associated with genus *Oxalobacter***

| **SNP** | **A1** | **A2** | **EAF** | **Beta** | **SE** | **N** | ***P*-value** |
| --- | --- | --- | --- | --- | --- | --- | --- |
| rs4428215^a^ | A | G | 0.755 | -0.005 | 0.003 | 268795 | 0.088 |
| rs736744 | C | T | 0.432 | -0.002 | 0.003 | 262880 | 0.423 |
| rs6000536 | T | C | 0.773 | -0.002 | 0.003 | 259212 | 0.610 |
| rs36057338 | T | G | 0.929 | 0.004 | 0.005 | 262446 | 0.429 |
| rs6071435 | A | T | 0.624 | -0.006 | 0.003 | 267796 | 0.043 |
| rs12002250 | C | A | 0.945 | 0.001 | 0.006 | 256693 | 0.814 |
| rs1569853 | C | T | 0.858 | -0.001 | 0.004 | 265825 | 0.770 |
| rs11108500^a^ | G | A | 0.935 | 0.010 | 0.006 | 261350 | 0.085 |
| rs10464997 | A | G | 0.844 | 0.009 | 0.004 | 254596 | 0.022 |
| rs111966731 | C | T | 0.943 | 0.005 | 0.006 | 262429 | 0.414 |
| rs6993398 | A | G | 0.836 | 0.009 | 0.004 | 265500 | 0.020 |
| rs3862635 | T | C | 0.920 | -0.012 | 0.005 | 262809 | 0.020 |

SNP: single nucleotide polymorphism; A1: effect allele; A2: baseline allele; EAF: effect allele frequency; SE: standard error. ^a^ SNPs were removed in the MR analysis during the heterogeneity test via RadialMR.

**Supplementary Table 5. Summary information on genus *Fusicatenibacter* SNPs used as genetic instruments for the Mendelian randomization analyses**

| **SNP** | **A1** | **A2** | **EAF** | **Beta** | **SE** | **N** | ***P*-value** | **R^2^ (%)** | **F-statistics** |
| --- | --- | --- | --- | --- | --- | --- | --- | --- | --- |
| rs4378146 | A | C | 0.242 | -0.062 | 0.013 | 17384 | 7.20E-07 | 0.139 | 24.3 |
| rs62353480 | A | G | 0.171 | -0.070 | 0.015 | 16119 | 1.57E-06 | 0.139 | 22.5 |
| rs704418 | T | C | 0.186 | 0.074 | 0.015 | 16626 | 7.77E-07 | 0.165 | 27.5 |
| rs2132128 | G | A | 0.165 | -0.077 | 0.016 | 17370 | 1.08E-06 | 0.164 | 28.6 |
| rs206581 | A | G | 0.209 | -0.057 | 0.013 | 17384 | 8.96E-06 | 0.107 | 18.6 |
| rs62187631 | T | C | 0.140 | -0.071 | 0.016 | 16522 | 4.55E-06 | 0.122 | 20.1 |
| rs2025938 | G | A | 0.077 | -0.097 | 0.021 | 15626 | 2.99E-06 | 0.132 | 20.7 |
| rs3303 | T | C | 0.077 | -0.095 | 0.020 | 16877 | 3.94E-06 | 0.129 | 21.7 |
| rs2039204 | T | A | 0.431 | -0.050 | 0.011 | 17383 | 3.94E-06 | 0.121 | 21.1 |
| rs8028026 | A | G | 0.091 | -0.079 | 0.018 | 16522 | 8.06E-06 | 0.104 | 17.2 |
| rs1864685 | A | C | 0.465 | -0.049 | 0.011 | 17374 | 4.96E-06 | 0.122 | 21.2 |
| rs792108 | T | C | 0.354 | -0.051 | 0.011 | 16978 | 8.50E-06 | 0.118 | 20.1 |
| rs60254196 | A | G | 0.541 | -0.049 | 0.011 | 16978 | 5.47E-06 | 0.120 | 20.5 |
| rs9905659 | G | A | 0.188 | -0.062 | 0.014 | 17384 | 7.31E-06 | 0.116 | 20.2 |
| rs8063430 | T | C | 0.060 | -0.104 | 0.022 | 15552 | 4.93E-06 | 0.121 | 18.9 |
| rs6515626 | G | A | 0.062 | 0.142 | 0.031 | 7796 | 7.29E-06 | 0.232 | 18.1 |
| rs10439674 | A | G | 0.202 | -0.057 | 0.013 | 17381 | 7.68E-06 | 0.105 | 18.3 |
| rs167879 | C | T | 0.167 | -0.066 | 0.015 | 16542 | 5.87E-06 | 0.121 | 20.0 |
| rs73103914 | A | G | 0.178 | -0.060 | 0.013 | 17384 | 8.30E-06 | 0.104 | 18.2 |

SNP: single nucleotide polymorphism; A1: effect allele; A2: baseline allele; EAF: effect allele frequency; SE: standard error; R^2^: Explained incremental variance in the phenotype.

**Supplementary Table 6. Summary information on intelligence for the 19 genome-wide significant SNPs associated with genus *Fusicatenibacter***

| **SNP ^a^** | **A1** | **A2** | **EAF** | **Beta** | **SE** | **N** | ***P*-value** |
| --- | --- | --- | --- | --- | --- | --- | --- |
| rs4378146 | C | A | 0.755 | 1.05E-4 | 0.003 | 267721 | 0.973 |
| rs206581 | G | A | 0.775 | 0.003 | 0.003 | 259983 | 0.316 |
| rs62187631 | C | T | 0.862 | 0.005 | 0.004 | 264136 | 0.193 |
| rs2025938 | A | G | 0.926 | 0.001 | 0.005 | 262568 | 0.845 |
| rs3303 | C | T | 0.926 | 0.006 | 0.005 | 244995 | 0.247 |
| rs8028026^b^ | A | G | 0.101 | 0.011 | 0.005 | 267517 | 0.012 |
| rs1864685 | C | A | 0.555 | 0.004 | 0.003 | 264902 | 0.177 |
| rs792108 | T | C | 0.354 | -0.005 | 0.003 | 268934 | 0.102 |
| rs60254196 | A | G | 0.516 | -0.001 | 0.003 | 255518 | 0.633 |
| rs9905659 | A | G | 0.803 | 0.005 | 0.003 | 261799 | 0.134 |
| rs8063430 | C | T | 0.933 | 0.008 | 0.006 | 259258 | 0.133 |
| rs6515626 | A | G | 0.936 | -0.001 | 0.006 | 259726 | 0.824 |
| rs10439674 | G | A | 0.780 | 0.002 | 0.003 | 264816 | 0.621 |
| rs167879 | C | T | 0.168 | -0.001 | 0.004 | 263591 | 0.739 |
| rs73103914 | G | A | 0.807 | 0.008 | 0.003 | 264791 | 0.026 |

SNP: single nucleotide polymorphism; A1: effect allele; A2: baseline allele; EAF: effect allele frequency; SE: standard error. ^a^ rs2039204, rs2132128, rs62353480 and rs704418 were not available in the outcome GWAS, and LDlink didn’t detect closely related SNP (r^2^ > 0.8) associated with the *genus Fusicatenibacter* (*P* < 1×10^-5^) and available in the outcome GWAS. Thus, these four SNPs were removed in the MR analysis. ^b^ SNPs were removed in the MR analysis during the heterogeneity test via RadialMR.

**Supplementary Table 7. Pleiotropy assessment results for the identified gut microbiota**

| **Method** | ***Oxalobacter* on intelligence** | ***Fusicatenibacter* on intelligence** | **Intelligence on *Oxalobacter*** | **Intelligence on *Fusicatenibacter*** |
| --- | --- | --- | --- | --- |
| MR Egger intercept | -0.005 | 0.003 | -0.004 | 1.83E-6 |
| MR Egger intercept 95%CI | -0.015 to 0.006 | -0.003 to 0.010 | -0.023 to 0.014 | -0.010 to 0.010 |
| MR Egger intercept *P* | 0.414 | 0.341 | 0.662 | 1 |
| MRPRESSO Global Test RSSobs | 10.03 | 9.29 | 109.98 | 130.86 |
| MRPRESSO Global Test *P* | 0.537 | 0.877 | 0.997 | 0.950 |
| Cochran’s Q | 8.25 | 7.86 | 108.47 | 129.12 |
| Cochran’s Q *P* | 0.509 | 0.853 | 0.997 | 0.928 |
| Rucker’s Qʹ | 7.51 | 6.88 | 108.28 | 129.12 |
| Rucker’s Qʹ *P* | 0.483 | 0.866 | 0.997 | 0.920 |
| Q-Q' | 0.74 | 0.98 | 0.19 | 0 |
| Q-Q' *P* | 0.389 | 0.322 | 0.661 | 1 |

**Supplementary Table 8. Summary information on intelligence SNPs used as genetic instruments used for the Mendelian randomization analyses**

| **SNP** | **A1** | **A2** | **EAF** | **Beta** | **SE** | **N** | ***P* value** | **R^2^ (%)** | **F-statistics** |
| --- | --- | --- | --- | --- | --- | --- | --- | --- | --- |
| rs10917152 | C | T | 0.869 | -0.024 | 0.004 | 267975 | 2.23E-09 | 0.013 | 35.8 |
| rs7518151 | T | C | 0.320 | -0.017 | 0.003 | 264585 | 1.03E-08 | 0.012 | 32.8 |
| rs12035012 | C | A | 0.783 | 0.027 | 0.003 | 268142 | 3.68E-16 | 0.025 | 66.4 |
| rs11210871 | C | G | 0.320 | -0.017 | 0.003 | 268102 | 3.08E-09 | 0.013 | 35.1 |
| rs1831539 | T | C | 0.543 | -0.017 | 0.003 | 264014 | 4.72E-10 | 0.015 | 38.8 |
| rs2420551 | T | A | 0.113 | 0.029 | 0.004 | 264461 | 4.60E-11 | 0.016 | 43.3 |
| rs3128341 | T | C | 0.198 | -0.032 | 0.003 | 269451 | 1.63E-20 | 0.032 | 86.2 |
| rs6668048 | C | T | 0.528 | 0.021 | 0.003 | 268318 | 4.24E-15 | 0.023 | 61.6 |
| rs1473474 | T | C | 0.472 | 0.016 | 0.003 | 267657 | 1.33E-08 | 0.012 | 32.3 |
| rs12026245 | G | A | 0.504 | 0.018 | 0.003 | 268403 | 6.05E-11 | 0.016 | 42.8 |
| rs600806 | G | A | 0.274 | 0.019 | 0.003 | 265886 | 3.57E-10 | 0.015 | 39.3 |
| rs112780312 | G | A | 0.716 | 0.018 | 0.003 | 256189 | 3.66E-09 | 0.014 | 34.8 |
| rs199928 | T | C | 0.175 | 0.020 | 0.004 | 264195 | 3.15E-08 | 0.012 | 30.6 |
| rs2678210 | T | C | 0.714 | 0.019 | 0.003 | 263842 | 6.97E-10 | 0.014 | 38.0 |
| rs10779271 | A | G | 0.681 | 0.016 | 0.003 | 269057 | 2.17E-08 | 0.012 | 31.3 |
| rs12470949 | T | C | 0.284 | -0.017 | 0.003 | 269277 | 1.32E-08 | 0.012 | 32.3 |
| rs967569 | C | T | 0.326 | 0.018 | 0.003 | 265548 | 8.21E-10 | 0.014 | 37.7 |
| rs2955280 | C | T | 0.472 | 0.015 | 0.003 | 268351 | 4.90E-08 | 0.011 | 29.8 |
| rs58593843 | G | A | 0.903 | 0.028 | 0.005 | 263810 | 2.67E-09 | 0.013 | 35.4 |
| rs10189857 | A | G | 0.563 | 0.019 | 0.003 | 268710 | 4.91E-12 | 0.018 | 47.7 |
| rs13395129 | G | C | 0.693 | -0.017 | 0.003 | 261678 | 1.83E-08 | 0.012 | 31.7 |
| rs4852252 | T | C | 0.450 | -0.021 | 0.003 | 267580 | 3.84E-14 | 0.021 | 57.2 |
| rs11898362 | G | A | 0.697 | 0.018 | 0.003 | 261477 | 2.25E-09 | 0.014 | 35.7 |
| rs11678106 | C | T | 0.503 | -0.016 | 0.003 | 265394 | 4.62E-09 | 0.013 | 34.3 |
| rs2309812 | C | T | 0.642 | -0.023 | 0.003 | 268685 | 9.95E-16 | 0.024 | 64.4 |
| rs60262711 | C | T | 0.615 | -0.016 | 0.003 | 264536 | 1.65E-08 | 0.012 | 31.9 |
| rs2558096 | T | G | 0.420 | -0.016 | 0.003 | 266795 | 1.74E-08 | 0.012 | 31.8 |
| rs10189912 | A | G | 0.646 | -0.019 | 0.003 | 268761 | 1.22E-11 | 0.017 | 45.9 |
| rs297578 | G | A | 0.294 | -0.018 | 0.003 | 266530 | 1.73E-09 | 0.014 | 36.3 |
| rs2268894 | C | T | 0.453 | -0.021 | 0.003 | 267027 | 3.98E-14 | 0.021 | 57.2 |
| rs6432749 | G | A | 0.724 | 0.017 | 0.003 | 262784 | 1.74E-08 | 0.012 | 31.8 |
| rs62181012 | T | C | 0.808 | 0.021 | 0.004 | 261314 | 1.73E-09 | 0.014 | 36.3 |
| rs62198803 | G | A | 0.764 | -0.019 | 0.003 | 266194 | 3.40E-09 | 0.013 | 34.9 |
| rs7573001 | G | C | 0.620 | 0.016 | 0.003 | 259519 | 1.32E-08 | 0.012 | 32.3 |
| rs2007176 | T | C | 0.531 | 0.015 | 0.003 | 262025 | 2.64E-08 | 0.012 | 31.0 |
| rs35731967 | T | C | 0.820 | 0.022 | 0.004 | 252991 | 2.38E-09 | 0.014 | 35.6 |
| rs13024268 | G | A | 0.614 | 0.017 | 0.003 | 254536 | 7.15E-09 | 0.013 | 33.5 |
| rs6550835 | G | A | 0.674 | 0.025 | 0.003 | 265082 | 2.44E-17 | 0.027 | 71.8 |
| rs1589652 | A | G | 0.443 | 0.017 | 0.003 | 266070 | 5.82E-10 | 0.014 | 38.4 |
| rs2352974 | C | T | 0.538 | 0.031 | 0.003 | 265625 | 3.69E-29 | 0.047 | 125.6 |
| rs7640196 | C | T | 0.751 | 0.017 | 0.003 | 269021 | 3.15E-08 | 0.011 | 30.6 |
| rs11720523 | C | A | 0.590 | -0.018 | 0.003 | 268784 | 3.89E-11 | 0.016 | 43.7 |
| rs6770622 | G | A | 0.958 | 0.045 | 0.007 | 265221 | 5.76E-11 | 0.016 | 42.9 |
| rs7652296 | A | G | 0.610 | 0.017 | 0.003 | 268118 | 3.51E-09 | 0.013 | 34.9 |
| rs3860537 | T | C | 0.210 | 0.019 | 0.003 | 260387 | 2.99E-08 | 0.012 | 30.7 |
| rs13071190 | T | C | 0.667 | 0.018 | 0.003 | 264545 | 5.55E-10 | 0.015 | 38.5 |
| rs59142272 | G | A | 0.831 | -0.023 | 0.004 | 261961 | 7.32E-10 | 0.014 | 37.9 |
| rs12646225 | C | T | 0.880 | -0.025 | 0.004 | 266694 | 2.51E-09 | 0.013 | 35.5 |
| rs4484297 | G | C | 0.749 | -0.018 | 0.003 | 266417 | 7.45E-09 | 0.013 | 33.4 |
| rs144246 | G | A | 0.630 | -0.015 | 0.003 | 266008 | 4.91E-08 | 0.011 | 29.8 |
| rs34811474 | G | A | 0.802 | -0.029 | 0.004 | 243818 | 7.15E-16 | 0.027 | 65.1 |
| rs67482514 | C | G | 0.756 | -0.018 | 0.003 | 259981 | 3.21E-08 | 0.012 | 30.6 |
| rs6819372 | A | G | 0.493 | -0.020 | 0.003 | 268631 | 4.02E-13 | 0.020 | 52.6 |
| rs1972860 | G | A | 0.679 | 0.018 | 0.003 | 267176 | 2.09E-09 | 0.013 | 35.9 |
| rs17199964 | G | A | 0.938 | 0.039 | 0.006 | 264922 | 6.67E-12 | 0.018 | 47.1 |
| rs2726491 | G | A | 0.650 | 0.028 | 0.003 | 269092 | 4.17E-23 | 0.036 | 98.0 |
| rs6535809 | A | G | 0.513 | 0.020 | 0.003 | 267680 | 6.65E-13 | 0.019 | 51.6 |
| rs1840847 | G | A | 0.650 | -0.016 | 0.003 | 264327 | 1.44E-08 | 0.012 | 32.1 |
| rs75973558 | A | G | 0.885 | 0.026 | 0.004 | 245527 | 9.42E-09 | 0.013 | 33.0 |
| rs34426618 | C | T | 0.815 | 0.020 | 0.004 | 264140 | 2.16E-08 | 0.012 | 31.3 |
| rs36033 | T | C | 0.582 | 0.016 | 0.003 | 264416 | 1.02E-08 | 0.012 | 32.8 |
| rs1812587 | G | T | 0.517 | 0.017 | 0.003 | 261560 | 3.68E-10 | 0.015 | 39.3 |
| rs80170948 | T | G | 0.961 | 0.045 | 0.007 | 246519 | 7.69E-10 | 0.015 | 37.8 |
| rs34316 | A | C | 0.435 | 0.021 | 0.003 | 265617 | 2.82E-14 | 0.022 | 57.9 |
| rs166820 | G | A | 0.826 | -0.024 | 0.004 | 269027 | 1.37E-11 | 0.017 | 45.7 |
| rs55763037 | A | G | 0.781 | 0.018 | 0.003 | 263387 | 3.74E-08 | 0.011 | 30.3 |
| rs12187824 | C | A | 0.515 | -0.015 | 0.003 | 265904 | 2.43E-08 | 0.012 | 31.1 |
| rs1145123 | T | C | 0.516 | 0.021 | 0.003 | 260587 | 1.20E-13 | 0.021 | 55.0 |
| rs405321 | G | A | 0.699 | 0.016 | 0.003 | 268718 | 3.32E-08 | 0.011 | 30.5 |
| rs4463213 | G | A | 0.479 | -0.019 | 0.003 | 268325 | 3.00E-12 | 0.018 | 48.7 |
| rs31768 | A | T | 0.287 | 0.018 | 0.003 | 262143 | 2.65E-09 | 0.014 | 35.4 |
| rs6860963 | C | T | 0.810 | -0.020 | 0.003 | 268942 | 5.57E-09 | 0.013 | 34.0 |
| rs2450333 | G | A | 0.505 | 0.019 | 0.003 | 255271 | 1.73E-11 | 0.018 | 45.3 |
| rs9503599 | T | C | 0.563 | -0.017 | 0.003 | 261955 | 8.05E-10 | 0.014 | 37.7 |
| rs566237 | A | G | 0.683 | -0.019 | 0.003 | 268091 | 1.82E-10 | 0.015 | 40.7 |
| rs6903716 | A | G | 0.700 | 0.018 | 0.003 | 268979 | 2.39E-09 | 0.013 | 35.6 |
| rs35433030 | G | A | 0.917 | -0.027 | 0.005 | 265521 | 4.67E-08 | 0.011 | 29.8 |
| rs1280049 | A | C | 0.475 | 0.015 | 0.003 | 269184 | 3.92E-08 | 0.011 | 30.2 |
| rs12190777 | A | G | 0.722 | 0.017 | 0.003 | 260594 | 3.63E-08 | 0.012 | 30.3 |
| rs1906252 | C | A | 0.534 | -0.032 | 0.003 | 267212 | 7.48E-31 | 0.050 | 133.4 |
| rs9384679 | C | T | 0.601 | 0.027 | 0.003 | 269078 | 7.94E-22 | 0.034 | 92.2 |
| rs13212044 | G | T | 0.769 | 0.018 | 0.003 | 268153 | 1.46E-08 | 0.012 | 32.1 |
| rs287879 | A | G | 0.731 | -0.019 | 0.003 | 268781 | 8.47E-10 | 0.014 | 37.7 |
| rs4725065 | A | G | 0.518 | -0.017 | 0.003 | 267463 | 1.52E-09 | 0.014 | 36.5 |
| rs115064 | T | C | 0.612 | 0.016 | 0.003 | 265957 | 1.07E-08 | 0.012 | 32.7 |
| rs1376289 | G | C | 0.678 | 0.016 | 0.003 | 268815 | 4.07E-08 | 0.011 | 30.1 |
| rs799444 | T | C | 0.451 | 0.018 | 0.003 | 265222 | 2.48E-11 | 0.017 | 44.5 |
| rs13223152 | A | G | 0.591 | 0.018 | 0.003 | 266940 | 2.34E-10 | 0.015 | 40.2 |
| rs56150095 | C | A | 0.468 | 0.022 | 0.003 | 266047 | 1.28E-15 | 0.024 | 64.0 |
| rs12535854 | C | G | 0.336 | -0.018 | 0.003 | 256877 | 6.73E-10 | 0.015 | 38.1 |
| rs4731392 | A | G | 0.692 | -0.022 | 0.003 | 265178 | 2.69E-13 | 0.020 | 53.4 |
| rs1362739 | C | A | 0.531 | -0.021 | 0.003 | 268624 | 1.83E-14 | 0.022 | 58.7 |
| rs13253386 | T | G | 0.540 | -0.020 | 0.003 | 266502 | 2.37E-13 | 0.020 | 53.7 |
| rs10954779 | C | T | 0.444 | 0.016 | 0.003 | 265561 | 3.04E-09 | 0.013 | 35.2 |
| rs13276212 | G | T | 0.517 | -0.015 | 0.003 | 263852 | 4.48E-08 | 0.011 | 29.9 |
| rs2920940 | T | C | 0.231 | -0.025 | 0.003 | 266393 | 2.76E-14 | 0.022 | 57.9 |
| rs2111490 | A | G | 0.465 | 0.015 | 0.003 | 265184 | 1.83E-08 | 0.012 | 31.7 |
| rs7357604 | A | G | 0.622 | 0.016 | 0.003 | 267314 | 2.56E-08 | 0.012 | 31.0 |
| rs4976976 | G | A | 0.581 | -0.017 | 0.003 | 266185 | 4.53E-10 | 0.015 | 38.9 |
| rs2721173 | C | T | 0.510 | 0.016 | 0.003 | 267693 | 2.89E-09 | 0.013 | 35.3 |
| rs11793831 | G | T | 0.592 | -0.028 | 0.003 | 263167 | 3.25E-23 | 0.037 | 98.5 |
| rs702222 | C | T | 0.644 | 0.020 | 0.003 | 264469 | 5.02E-12 | 0.018 | 47.7 |
| rs28620532 | A | G | 0.644 | -0.016 | 0.003 | 261069 | 1.51E-08 | 0.012 | 32.0 |
| rs913264 | C | T | 0.713 | -0.020 | 0.003 | 267060 | 7.09E-11 | 0.016 | 42.5 |
| rs2987390 | C | G | 0.733 | -0.018 | 0.003 | 262158 | 1.19E-08 | 0.012 | 32.5 |
| rs7069887 | A | C | 0.852 | 0.023 | 0.004 | 261247 | 7.44E-09 | 0.013 | 33.4 |
| rs2393967 | A | C | 0.691 | -0.019 | 0.003 | 266989 | 2.70E-10 | 0.015 | 39.9 |
| rs1408579 | C | T | 0.554 | -0.016 | 0.003 | 267937 | 5.23E-09 | 0.013 | 34.1 |
| rs3740422 | G | C | 0.674 | 0.024 | 0.003 | 268246 | 1.25E-16 | 0.026 | 68.5 |
| rs35608616 | G | A | 0.669 | 0.018 | 0.003 | 261906 | 7.33E-10 | 0.014 | 37.9 |
| rs11605348 | G | A | 0.658 | 0.017 | 0.003 | 264878 | 9.73E-09 | 0.012 | 32.9 |
| rs7941785 | A | G | 0.369 | 0.016 | 0.003 | 265984 | 4.75E-08 | 0.011 | 29.8 |
| rs2373353 | A | G | 0.637 | -0.016 | 0.003 | 259547 | 1.56E-08 | 0.012 | 32.0 |
| rs2508713 | T | A | 0.638 | -0.017 | 0.003 | 268362 | 5.92E-09 | 0.013 | 33.9 |
| rs7116046 | C | T | 0.629 | -0.016 | 0.003 | 265132 | 3.27E-08 | 0.012 | 30.5 |
| rs2885208 | T | C | 0.808 | 0.019 | 0.003 | 268192 | 4.58E-08 | 0.011 | 29.9 |
| rs17128425 | T | A | 0.899 | -0.026 | 0.005 | 267721 | 1.87E-08 | 0.012 | 31.6 |
| rs329672 | C | T | 0.372 | -0.017 | 0.003 | 262794 | 1.00E-09 | 0.014 | 37.3 |
| rs55754731 | T | C | 0.831 | 0.021 | 0.004 | 264037 | 6.06E-09 | 0.013 | 33.8 |
| rs1054442 | A | C | 0.620 | -0.021 | 0.003 | 267489 | 2.52E-14 | 0.022 | 58.1 |
| rs1962047 | G | A | 0.638 | 0.020 | 0.003 | 264057 | 8.89E-12 | 0.018 | 46.6 |
| rs6539284 | T | C | 0.613 | -0.019 | 0.003 | 263597 | 5.56E-12 | 0.018 | 47.5 |
| rs7312919 | C | G | 0.663 | 0.018 | 0.003 | 263162 | 4.83E-10 | 0.015 | 38.8 |
| rs1727307 | A | G | 0.289 | 0.018 | 0.003 | 269374 | 3.10E-09 | 0.013 | 35.1 |
| rs9569206 | A | G | 0.629 | -0.015 | 0.003 | 268498 | 4.85E-08 | 0.011 | 29.8 |
| rs9316954 | C | G | 0.296 | -0.017 | 0.003 | 269324 | 6.05E-09 | 0.013 | 33.8 |
| rs9516855 | A | G | 0.947 | 0.033 | 0.006 | 266748 | 4.19E-08 | 0.011 | 30.1 |
| rs2478286 | G | C | 0.254 | 0.026 | 0.003 | 269582 | 1.64E-16 | 0.025 | 68.0 |
| rs8006700 | T | A | 0.321 | 0.018 | 0.003 | 267288 | 4.96E-10 | 0.014 | 38.7 |
| rs4981713 | T | G | 0.605 | 0.016 | 0.003 | 267334 | 7.23E-09 | 0.013 | 33.5 |
| rs2239647 | A | C | 0.448 | -0.021 | 0.003 | 264189 | 1.14E-13 | 0.021 | 55.1 |
| rs11623436 | C | T | 0.553 | 0.016 | 0.003 | 267439 | 9.70E-09 | 0.012 | 32.9 |
| rs12886584 | T | C | 0.823 | 0.021 | 0.004 | 268772 | 9.41E-09 | 0.012 | 33.0 |
| rs17106817 | T | C | 0.707 | 0.017 | 0.003 | 263620 | 2.26E-08 | 0.012 | 31.3 |
| rs1007934 | G | A | 0.620 | -0.016 | 0.003 | 269586 | 1.00E-08 | 0.012 | 32.8 |
| rs2071407 | T | C | 0.362 | -0.022 | 0.003 | 264658 | 1.52E-14 | 0.022 | 59.1 |
| rs11634187 | T | G | 0.850 | 0.022 | 0.004 | 264197 | 1.12E-08 | 0.012 | 32.6 |
| rs7172979 | G | T | 0.976 | -0.061 | 0.009 | 260362 | 2.47E-11 | 0.017 | 44.6 |
| rs72739469 | T | C | 0.931 | -0.034 | 0.006 | 244196 | 1.18E-09 | 0.015 | 37.0 |
| rs8025964 | G | A | 0.535 | -0.017 | 0.003 | 265960 | 5.78E-10 | 0.014 | 38.4 |
| rs1369429 | T | C | 0.348 | 0.018 | 0.003 | 262697 | 1.15E-09 | 0.014 | 37.1 |
| rs11076962 | T | C | 0.717 | 0.017 | 0.003 | 266051 | 2.57E-08 | 0.012 | 31.0 |
| rs11646221 | G | T | 0.444 | -0.018 | 0.003 | 263669 | 1.57E-10 | 0.016 | 40.9 |
| rs2457192 | C | A | 0.277 | 0.020 | 0.003 | 254769 | 2.84E-10 | 0.016 | 39.8 |
| rs72768642 | T | C | 0.930 | -0.031 | 0.005 | 263505 | 1.46E-08 | 0.012 | 32.1 |
| rs2008514 | G | A | 0.615 | 0.029 | 0.003 | 269455 | 1.25E-24 | 0.039 | 105.0 |
| rs2647995 | T | C | 0.705 | -0.020 | 0.003 | 259642 | 8.68E-11 | 0.016 | 42.1 |
| rs8054299 | C | G | 0.682 | -0.023 | 0.003 | 269270 | 3.84E-15 | 0.023 | 61.8 |
| rs9888986 | G | A | 0.884 | 0.024 | 0.004 | 267786 | 3.52E-08 | 0.011 | 30.4 |
| rs8051038 | G | A | 0.251 | -0.019 | 0.003 | 269045 | 1.78E-09 | 0.013 | 36.2 |
| rs2285640 | G | A | 0.456 | -0.018 | 0.003 | 263690 | 2.38E-10 | 0.015 | 40.1 |
| rs4793161 | A | G | 0.232 | -0.018 | 0.003 | 265288 | 4.97E-08 | 0.011 | 29.7 |
| rs17698176 | T | G | 0.802 | -0.020 | 0.004 | 247702 | 1.70E-08 | 0.013 | 31.8 |
| rs11079849 | C | T | 0.687 | -0.017 | 0.003 | 265226 | 2.26E-08 | 0.012 | 31.3 |
| rs66954617 | A | G | 0.374 | -0.021 | 0.003 | 265761 | 1.72E-13 | 0.020 | 54.3 |
| rs6508220 | A | G | 0.503 | -0.023 | 0.003 | 266839 | 9.56E-17 | 0.026 | 69.1 |
| rs17002025 | G | A | 0.878 | -0.026 | 0.004 | 256305 | 1.89E-09 | 0.014 | 36.1 |
| rs2072490 | C | T | 0.491 | -0.017 | 0.003 | 265518 | 5.93E-10 | 0.014 | 38.3 |
| rs7248006 | T | C | 0.388 | -0.019 | 0.003 | 264719 | 1.05E-11 | 0.017 | 46.2 |
| rs144026674 | C | T | 0.962 | -0.041 | 0.007 | 247977 | 3.19E-08 | 0.012 | 30.6 |
| rs889169 | G | A | 0.399 | -0.016 | 0.003 | 249200 | 2.75E-08 | 0.012 | 30.9 |
| rs73068339 | G | C | 0.717 | -0.019 | 0.003 | 265789 | 5.96E-10 | 0.014 | 38.3 |
| rs78084033 | A | C | 0.866 | -0.023 | 0.004 | 262111 | 1.62E-08 | 0.012 | 31.9 |
| rs6019535 | G | A | 0.695 | -0.025 | 0.003 | 266115 | 3.28E-17 | 0.027 | 71.2 |
| rs2836921 | G | A | 0.688 | -0.020 | 0.003 | 265147 | 6.54E-12 | 0.018 | 47.2 |
| rs5753383 | G | A | 0.679 | -0.016 | 0.003 | 269175 | 5.00E-08 | 0.011 | 29.7 |
| rs5750830 | C | A | 0.259 | -0.023 | 0.003 | 266749 | 2.46E-13 | 0.020 | 53.6 |
| rs4821995 | A | G | 0.344 | 0.016 | 0.003 | 267995 | 2.62E-08 | 0.012 | 31.0 |

SNP: single nucleotide polymorphism; A1: effect allele; A2: baseline allele; EAF: effect allele frequency; SE: standard error; R^2^: Explained incremental variance in the phenotype.

**Supplementary Table 9. Summary information on genus *Oxalobacter* and genus *Fusicatenibacter* for the 168 genome-wide significant SNPs associated with intelligence**

| **SNP ^a^** | **A1** | **A2** | **EAF** | **genus *Oxalobacter*** | | | **genus *Fusicatenibacter*** | | |
| --- | --- | --- | --- | --- | --- | --- | --- | --- | --- |
|  |  |  |  | **Beta** | **SE** | ***P*-value** | **Beta** | **SE** | ***P*-value** |
| rs10917152 | T | C | 0.148 | -0.003 | 0.031 | 0.985 | -0.010 | 0.016 | 0.579 |
| rs7518151 | T | C | 0.291 | 0.037 | 0.023 | 0.099 | -0.011 | 0.012 | 0.336 |
| rs12035012 | A | C | 0.214 | -0.027 | 0.025 | 0.278 | -0.010 | 0.013 | 0.392 |
| rs11210871 | C | G | 0.331 | 0.034 | 0.023 | 0.126 | 0.019 | 0.012 | 0.097 |
| rs1831539 | C | T | 0.461 | -0.012 | 0.021 | 0.565 | 0.014 | 0.011 | 0.193 |
| rs2420551 | T | A | 0.104 | 0.021 | 0.035 | 0.553 | 0.002 | 0.018 | 0.944 |
| rs3128341^c^ | T | C | 0.184 | 0.030 | 0.025 | 0.279 | 0.028 | 0.013 | 0.033 |
| rs6668048 | C | T | 0.530 | -0.021 | 0.021 | 0.286 | 0.012 | 0.011 | 0.350 |
| rs1473474 | T | C | 0.459 | -0.013 | 0.021 | 0.523 | -0.001 | 0.011 | 0.938 |
| rs12026245 | G | A | 0.522 | -0.025 | 0.021 | 0.274 | -0.009 | 0.011 | 0.374 |
| rs600806 | G | A | 0.271 | 0.002 | 0.024 | 0.920 | 0.011 | 0.012 | 0.398 |
| rs112780312^b^ | A | G | 0.277 | 0.048 | 0.024 | 0.041 | 0.021 | 0.012 | 0.073 |
| rs199928 | T | C | 0.194 | 0.012 | 0.028 | 0.664 | -0.010 | 0.014 | 0.498 |
| rs2678210 | C | T | 0.291 | 0.019 | 0.023 | 0.399 | 0.003 | 0.012 | 0.817 |
| rs10779271 | G | A | 0.303 | 0.018 | 0.022 | 0.453 | -0.001 | 0.012 | 0.908 |
| rs12470949 | T | C | 0.277 | 0.004 | 0.023 | 0.805 | 0.002 | 0.012 | 0.885 |
| rs967569^b^ | C | T | 0.341 | 0.053 | 0.022 | 0.022 | 0.018 | 0.012 | 0.107 |
| rs2955280 | T | C | 0.500 | -0.018 | 0.021 | 0.435 | 0.004 | 0.011 | 0.697 |
| rs58593843 | A | G | 0.091 | -0.029 | 0.038 | 0.558 | -0.027 | 0.019 | 0.119 |
| rs10189857 | G | A | 0.421 | -0.004 | 0.021 | 0.848 | -0.014 | 0.011 | 0.185 |
| rs13395129^c^ | C | G | 0.291 | -0.020 | 0.022 | 0.400 | 0.029 | 0.012 | 0.016 |
| rs4852252^b^ | T | C | 0.455 | 0.041 | 0.021 | 0.052 | -0.002 | 0.011 | 0.845 |
| rs11898362 | A | G | 0.277 | 0.023 | 0.023 | 0.320 | 0.017 | 0.012 | 0.145 |
| rs11678106 | T | C | 0.513 | 0.022 | 0.021 | 0.287 | 0.009 | 0.011 | 0.422 |
| rs2309812^c^ | T | C | 0.389 | -0.005 | 0.021 | 0.820 | -0.028 | 0.011 | 0.013 |
| rs60262711 | T | C | 0.422 | 0.006 | 0.021 | 0.783 | 0.006 | 0.011 | 0.617 |
| rs2558096 | T | G | 0.431 | 0.018 | 0.021 | 0.407 | 0.006 | 0.011 | 0.554 |
| rs10189912 | G | A | 0.365 | 0.003 | 0.022 | 0.842 | 0.009 | 0.011 | 0.466 |
| rs297578 | G | A | 0.313 | -0.011 | 0.023 | 0.642 | -0.013 | 0.012 | 0.289 |
| rs2268894 | C | T | 0.440 | 0.001 | 0.021 | 0.960 | 0.001 | 0.011 | 0.922 |
| rs6432749 | A | G | 0.309 | -0.001 | 0.023 | 0.936 | -0.004 | 0.012 | 0.764 |
| rs62181012 | C | T | 0.190 | 0.017 | 0.026 | 0.474 | 0.018 | 0.014 | 0.205 |
| rs62198803 | A | G | 0.230 | -0.016 | 0.025 | 0.502 | 0.001 | 0.013 | 0.884 |
| rs7573001 | C | G | 0.419 | -0.025 | 0.021 | 0.226 | -0.018 | 0.011 | 0.090 |
| rs2007176 | C | T | 0.470 | 0.024 | 0.021 | 0.243 | 0.009 | 0.011 | 0.426 |
| rs35731967 | C | T | 0.195 | 0.027 | 0.028 | 0.314 | -0.008 | 0.015 | 0.498 |
| rs13024268 | A | G | 0.383 | -0.023 | 0.021 | 0.285 | 0.014 | 0.011 | 0.198 |
| rs6550835 | A | G | 0.302 | -0.025 | 0.022 | 0.300 | 0.000 | 0.012 | 0.914 |
| rs1589652 | A | G | 0.466 | 0.005 | 0.021 | 0.784 | 0.010 | 0.011 | 0.357 |
| rs2352974^c^ | T | C | 0.466 | 0.020 | 0.021 | 0.363 | 0.024 | 0.011 | 0.025 |
| rs7640196 | T | C | 0.199 | 0.021 | 0.025 | 0.382 | -0.005 | 0.013 | 0.713 |
| rs11720523^c^ | A | C | 0.436 | 0.023 | 0.021 | 0.282 | -0.030 | 0.011 | 0.007 |
| rs7652296 | G | A | 0.408 | 0.000 | 0.021 | 0.942 | 0.000 | 0.011 | 0.982 |
| rs3860537 | T | C | 0.227 | -0.005 | 0.026 | 0.833 | -0.004 | 0.014 | 0.785 |
| rs13071190 | C | T | 0.314 | 0.010 | 0.022 | 0.679 | 0.018 | 0.011 | 0.121 |
| rs59142272 | A | G | 0.183 | -0.036 | 0.027 | 0.215 | 0.003 | 0.014 | 0.834 |
| rs12646225 | T | C | 0.111 | -0.019 | 0.034 | 0.551 | -0.005 | 0.018 | 0.723 |
| rs4484297 | C | G | 0.241 | -0.028 | 0.024 | 0.276 | 0.005 | 0.012 | 0.667 |
| rs144246 | A | G | 0.355 | 0.022 | 0.022 | 0.353 | 0.017 | 0.011 | 0.126 |
| rs34811474 | A | G | 0.217 | -0.014 | 0.026 | 0.628 | -0.012 | 0.013 | 0.483 |
| rs67482514 | G | C | 0.243 | -0.029 | 0.025 | 0.276 | -0.015 | 0.013 | 0.221 |
| rs6819372 | G | A | 0.508 | 0.017 | 0.021 | 0.404 | 0.019 | 0.011 | 0.072 |
| rs1972860 | A | G | 0.286 | -0.004 | 0.022 | 0.848 | 0.001 | 0.011 | 0.942 |
| rs17199964 | A | G | 0.066 | -0.039 | 0.046 | 0.414 | -0.008 | 0.025 | 0.737 |
| rs2726491 | A | G | 0.371 | 0.016 | 0.022 | 0.487 | 0.020 | 0.011 | 0.084 |
| rs6535809 | G | A | 0.484 | -0.032 | 0.021 | 0.112 | -0.007 | 0.011 | 0.503 |
| rs1840847 | A | G | 0.332 | 0.016 | 0.022 | 0.481 | -0.010 | 0.011 | 0.369 |
| rs75973558 | G | A | 0.112 | 0.016 | 0.034 | 0.648 | -0.002 | 0.018 | 0.843 |
| rs34426618 | T | C | 0.144 | 0.000 | 0.027 | 0.993 | 0.006 | 0.014 | 0.687 |
| rs36033 | C | T | 0.429 | 0.035 | 0.021 | 0.101 | -0.001 | 0.011 | 0.913 |
| rs1812587 | T | G | 0.487 | -0.013 | 0.021 | 0.596 | -0.001 | 0.011 | 0.924 |
| rs34316 | A | C | 0.444 | 0.028 | 0.021 | 0.190 | 0.011 | 0.011 | 0.313 |
| rs166820 | A | G | 0.162 | 0.025 | 0.028 | 0.398 | -0.005 | 0.014 | 0.648 |
| rs55763037 | G | A | 0.203 | 0.045 | 0.024 | 0.068 | 0.017 | 0.012 | 0.228 |
| rs12187824 | C | A | 0.499 | 0.001 | 0.021 | 0.990 | 0.007 | 0.011 | 0.541 |
| rs1145123 | T | C | 0.516 | 0.018 | 0.021 | 0.416 | 0.003 | 0.011 | 0.730 |
| rs405321 | A | G | 0.347 | 0.010 | 0.023 | 0.657 | 0.011 | 0.012 | 0.355 |
| rs4463213 | G | A | 0.449 | -0.002 | 0.021 | 0.926 | -0.006 | 0.011 | 0.572 |
| rs31768 | A | T | 0.284 | 0.005 | 0.023 | 0.885 | -0.001 | 0.012 | 0.848 |
| rs6860963 | T | C | 0.206 | -0.017 | 0.026 | 0.497 | -0.022 | 0.013 | 0.077 |
| rs2450333 | G | A | 0.502 | 0.014 | 0.021 | 0.489 | 0.005 | 0.011 | 0.626 |
| rs9503599 | C | T | 0.434 | -0.005 | 0.021 | 0.789 | 0.002 | 0.011 | 0.836 |
| rs566237 | G | A | 0.319 | -0.008 | 0.022 | 0.731 | 0.022 | 0.012 | 0.051 |
| rs6903716 | G | A | 0.287 | -0.002 | 0.023 | 0.930 | -0.014 | 0.012 | 0.252 |
| rs35433030^b^ | A | G | 0.082 | -0.079 | 0.036 | 0.029 | 0.028 | 0.019 | 0.145 |
| rs1280049 | A | C | 0.478 | -0.007 | 0.021 | 0.749 | -0.001 | 0.011 | 0.941 |
| rs12190777 | G | A | 0.294 | -0.016 | 0.024 | 0.482 | 0.007 | 0.012 | 0.553 |
| rs1906252 | A | C | 0.507 | 0.033 | 0.021 | 0.121 | 0.003 | 0.011 | 0.815 |
| rs9384679 | T | C | 0.433 | 0.018 | 0.021 | 0.400 | 0.004 | 0.011 | 0.688 |
| rs13212044 | T | G | 0.242 | -0.008 | 0.025 | 0.711 | -0.021 | 0.013 | 0.113 |
| rs287879 | G | A | 0.288 | 0.009 | 0.023 | 0.727 | 0.014 | 0.012 | 0.236 |
| rs4725065 | G | A | 0.467 | -0.028 | 0.021 | 0.169 | -0.019 | 0.011 | 0.076 |
| rs115064 | C | T | 0.365 | -0.005 | 0.021 | 0.802 | 0.000 | 0.011 | 0.973 |
| rs1376289 | C | G | 0.323 | 0.008 | 0.022 | 0.737 | -0.008 | 0.012 | 0.504 |
| rs799444 | T | C | 0.466 | -0.005 | 0.021 | 0.807 | 0.011 | 0.011 | 0.330 |
| rs13223152 | G | A | 0.425 | -0.016 | 0.021 | 0.474 | 0.006 | 0.011 | 0.580 |
| rs56150095 | C | A | 0.426 | 0.020 | 0.021 | 0.355 | 0.011 | 0.011 | 0.320 |
| rs12535854 | C | G | 0.346 | -0.041 | 0.022 | 0.061 | 0.006 | 0.011 | 0.597 |
| rs4731392 | G | A | 0.310 | 0.031 | 0.023 | 0.172 | 0.020 | 0.012 | 0.111 |
| rs1362739 | A | C | 0.475 | 0.011 | 0.021 | 0.606 | 0.000 | 0.011 | 0.997 |
| rs13253386 | G | T | 0.479 | 0.005 | 0.021 | 0.801 | 0.001 | 0.011 | 0.857 |
| rs10954779 | C | T | 0.434 | 0.023 | 0.021 | 0.290 | 0.000 | 0.011 | 0.991 |
| rs13276212 | G | T | 0.496 | 0.010 | 0.021 | 0.642 | 0.000 | 0.011 | 0.929 |
| rs2920940 | T | C | 0.232 | -0.014 | 0.024 | 0.547 | 0.005 | 0.013 | 0.689 |
| rs2111490 | A | G | 0.439 | 0.005 | 0.021 | 0.801 | 0.018 | 0.011 | 0.091 |
| rs7357604 | G | A | 0.368 | -0.009 | 0.021 | 0.691 | 0.011 | 0.011 | 0.288 |
| rs4976976 | A | G | 0.407 | 0.004 | 0.021 | 0.877 | 0.003 | 0.011 | 0.794 |
| rs2721173^b^ | T | C | 0.461 | -0.044 | 0.021 | 0.039 | -0.009 | 0.011 | 0.432 |
| rs11793831^c^ | T | G | 0.432 | 0.009 | 0.021 | 0.676 | 0.020 | 0.011 | 0.067 |
| rs702222 | T | C | 0.358 | 0.024 | 0.022 | 0.263 | -0.016 | 0.011 | 0.154 |
| rs28620532 | G | A | 0.338 | -0.003 | 0.022 | 0.879 | 0.000 | 0.011 | 0.977 |
| rs913264 | T | C | 0.270 | 0.016 | 0.023 | 0.491 | -0.008 | 0.012 | 0.507 |
| rs2987390 | G | C | 0.300 | 0.008 | 0.024 | 0.804 | -0.016 | 0.012 | 0.219 |
| rs7069887 | C | A | 0.137 | -0.006 | 0.029 | 0.903 | -0.007 | 0.015 | 0.660 |
| rs2393967 | C | A | 0.336 | 0.015 | 0.022 | 0.470 | 0.001 | 0.012 | 0.942 |
| rs1408579 | T | C | 0.481 | -0.010 | 0.021 | 0.665 | -0.008 | 0.011 | 0.513 |
| rs3740422 | C | G | 0.344 | 0.028 | 0.022 | 0.208 | 0.007 | 0.011 | 0.505 |
| rs35608616^c^ | A | G | 0.317 | -0.016 | 0.022 | 0.504 | 0.034 | 0.011 | 0.004 |
| rs11605348 | A | G | 0.354 | -0.007 | 0.022 | 0.736 | 0.012 | 0.011 | 0.306 |
| rs7941785 | A | G | 0.350 | 0.017 | 0.022 | 0.423 | -0.001 | 0.011 | 0.973 |
| rs2373353^b^ | G | A | 0.354 | 0.054 | 0.022 | 0.012 | 0.012 | 0.011 | 0.307 |
| rs2508713^b^ | A | T | 0.374 | -0.046 | 0.021 | 0.033 | 0.000 | 0.011 | 0.963 |
| rs7116046^b^ | T | C | 0.390 | -0.047 | 0.021 | 0.033 | 0.009 | 0.011 | 0.405 |
| rs2885208 | C | T | 0.205 | 0.028 | 0.027 | 0.301 | -0.010 | 0.014 | 0.433 |
| rs17128425 | A | T | 0.108 | -0.021 | 0.035 | 0.617 | -0.006 | 0.018 | 0.757 |
| rs329672 | C | T | 0.389 | 0.028 | 0.022 | 0.188 | -0.010 | 0.011 | 0.358 |
| rs55754731 | C | T | 0.196 | -0.027 | 0.028 | 0.350 | 0.002 | 0.015 | 0.943 |
| rs1054442 | C | A | 0.361 | 0.014 | 0.021 | 0.514 | -0.010 | 0.011 | 0.389 |
| rs1962047 | A | G | 0.373 | 0.034 | 0.021 | 0.115 | -0.017 | 0.011 | 0.132 |
| rs6539284 | C | T | 0.419 | 0.025 | 0.021 | 0.236 | -0.006 | 0.011 | 0.632 |
| rs7312919 | G | C | 0.371 | 0.013 | 0.022 | 0.543 | 0.018 | 0.011 | 0.103 |
| rs1727307 | A | G | 0.307 | -0.028 | 0.023 | 0.238 | -0.001 | 0.012 | 0.960 |
| rs9569206 | G | A | 0.342 | -0.008 | 0.022 | 0.700 | -0.009 | 0.011 | 0.440 |
| rs9316954 | C | G | 0.258 | -0.035 | 0.022 | 0.133 | -0.007 | 0.012 | 0.515 |
| rs9516855 | G | A | 0.050 | -0.029 | 0.050 | 0.577 | -0.029 | 0.025 | 0.335 |
| rs2478286 | G | C | 0.258 | 0.033 | 0.024 | 0.164 | -0.022 | 0.012 | 0.068 |
| rs8006700 | T | A | 0.309 | 0.018 | 0.023 | 0.425 | -0.002 | 0.012 | 0.841 |
| rs4981713 | G | T | 0.404 | -0.001 | 0.021 | 0.945 | -0.004 | 0.011 | 0.690 |
| rs2239647^b^ | A | C | 0.457 | -0.045 | 0.021 | 0.038 | 0.020 | 0.011 | 0.067 |
| rs11623436 | C | T | 0.563 | -0.032 | 0.021 | 0.134 | -0.008 | 0.011 | 0.484 |
| rs12886584 | C | T | 0.160 | 0.034 | 0.026 | 0.191 | 0.008 | 0.014 | 0.522 |
| rs17106817 | C | T | 0.282 | -0.009 | 0.023 | 0.721 | -0.013 | 0.012 | 0.331 |
| rs1007934 | A | G | 0.397 | 0.014 | 0.021 | 0.540 | 0.003 | 0.011 | 0.861 |
| rs2071407 | T | C | 0.372 | -0.016 | 0.022 | 0.499 | -0.014 | 0.011 | 0.198 |
| rs11634187^b^ | G | T | 0.150 | -0.071 | 0.028 | 0.011 | -0.003 | 0.015 | 0.825 |
| rs72739469 | C | T | 0.061 | -0.030 | 0.045 | 0.534 | 0.000 | 0.024 | 0.969 |
| rs8025964 | A | G | 0.481 | 0.018 | 0.021 | 0.367 | -0.010 | 0.011 | 0.359 |
| rs1369429 | T | C | 0.320 | -0.011 | 0.022 | 0.620 | -0.014 | 0.011 | 0.219 |
| rs11076962 | C | T | 0.284 | 0.011 | 0.023 | 0.627 | 0.018 | 0.012 | 0.147 |
| rs11646221 | G | T | 0.428 | -0.018 | 0.021 | 0.398 | 0.013 | 0.011 | 0.226 |
| rs2457192 | C | A | 0.280 | 0.015 | 0.023 | 0.501 | -0.007 | 0.012 | 0.532 |
| rs72768642 | C | T | 0.092 | 0.012 | 0.041 | 0.672 | -0.020 | 0.022 | 0.425 |
| rs2008514 | A | G | 0.331 | 0.000 | 0.021 | 0.985 | -0.003 | 0.011 | 0.873 |
| rs2647995 | C | T | 0.272 | -0.023 | 0.023 | 0.345 | 0.015 | 0.012 | 0.249 |
| rs8054299^c^ | G | C | 0.304 | -0.005 | 0.022 | 0.777 | -0.024 | 0.012 | 0.036 |
| rs9888986 | A | G | 0.116 | -0.004 | 0.033 | 0.817 | -0.004 | 0.017 | 0.962 |
| rs8051038 | G | A | 0.263 | 0.033 | 0.024 | 0.141 | -0.004 | 0.012 | 0.806 |
| rs2285640 | G | A | 0.448 | -0.014 | 0.021 | 0.494 | -0.014 | 0.011 | 0.196 |
| rs4793161 | A | G | 0.205 | -0.010 | 0.024 | 0.676 | -0.002 | 0.013 | 0.858 |
| rs17698176^c^ | G | T | 0.228 | 0.012 | 0.026 | 0.598 | -0.045 | 0.013 | 0.000 |
| rs11079849 | T | C | 0.290 | -0.030 | 0.023 | 0.185 | -0.007 | 0.012 | 0.532 |
| rs66954617 | A | G | 0.366 | 0.004 | 0.021 | 0.850 | -0.012 | 0.011 | 0.292 |
| rs6508220 | G | A | 0.499 | -0.006 | 0.021 | 0.786 | 0.018 | 0.011 | 0.099 |
| rs17002025 | A | G | 0.129 | 0.012 | 0.032 | 0.717 | -0.010 | 0.017 | 0.570 |
| rs2072490 | C | T | 0.498 | -0.029 | 0.021 | 0.159 | 0.009 | 0.011 | 0.404 |
| rs7248006 | T | C | 0.348 | -0.029 | 0.021 | 0.166 | 0.004 | 0.011 | 0.700 |
| rs889169 | G | A | 0.391 | 0.003 | 0.022 | 0.856 | -0.001 | 0.011 | 0.937 |
| rs73068339 | C | G | 0.276 | 0.008 | 0.023 | 0.676 | 0.009 | 0.012 | 0.442 |
| rs78084033 | C | A | 0.139 | 0.012 | 0.030 | 0.630 | 0.025 | 0.016 | 0.130 |
| rs6019535 | A | G | 0.295 | -0.032 | 0.022 | 0.154 | 0.000 | 0.012 | 0.946 |
| rs2836921 | A | G | 0.305 | 0.007 | 0.022 | 0.759 | 0.001 | 0.012 | 0.917 |
| rs5753383 | A | G | 0.310 | -0.023 | 0.022 | 0.315 | 0.008 | 0.012 | 0.471 |
| rs5750830 | C | A | 0.280 | 0.001 | 0.024 | 0.974 | 0.007 | 0.012 | 0.609 |
| rs4821995 | A | G | 0.334 | 0.013 | 0.022 | 0.541 | 0.010 | 0.011 | 0.385 |

SNP: single nucleotide polymorphism; A1: effect allele; A2: baseline allele; EAF: effect allele frequency; SE: standard error. ^a^ Four SNPs (rs6770622, rs80170948, rs7172979, and rs144026674) were not available in the GWAS of genus *Oxalobacter* and genus *Fusicatenibacter*, and LDlink didn’t detect closely related SNP (r^2^ > 0.8) associated with intelligence (*P* < 5×10^-8^) and available in the outcome GWAS. Thus, these SNPs were removed in the MR analysis. ^b^ SNPs were removed in the MR analysis for genus *Oxalobacter* during the heterogeneity test via RadialMR. ^c^ SNPs were removed in the MR analysis for genus *Fusicatenibacter* during the heterogeneity test via RadialMR.

**Supplementary Table 10. Significant MR results (*P* < 0.05) for the relationship between intelligence and gut microbiota**

| **Method** | **Number of SNPs** | ***F*-statistic** | **β (95% CI)** | ***P*-value** | ***q*-value** |
| --- | --- | --- | --- | --- | --- |
| Intelligence on genus *Ruminococcaceae UCG003* | | | | | |
| IVW | 159 | 42.3 | 0.175 (0.072 to 0.278) | 8.50×10^-4^ | 0.179 |
| MR RAPS |  |  | 0.177 (0.069 to 0.286) | 0.001 |  |
| Weighted Median |  |  | 0.134 (-0.016 to 0.284) | 0.080 |  |
| Weighted Mode |  |  | -0.145 (-0.640 to 0.350) | 0.562 |  |
| MR Egger |  |  | 0.156 (-0.386 to 0.698) | 0.571 |  |
| Intelligence on genus *Defluviitaleaceae UCG011* | | | | | |
| IVW | 154 | 41.9 | 0.208 (0.0693 to 0.347) | 0.003 | 0.347 |
| MR RAPS |  |  | 0.209 (0.063 to 0.356) | 0.005 |  |
| Weighted Median |  |  | 0.269 (0.077 to 0.461) | 0.006 |  |
| Weighted Mode |  |  | 0.250 (-0.326 to 0.826) | 0.392 |  |
| MR Egger |  |  | 0.937 (0.197 to 1.678) | 0.013 |  |
| Intelligence on genus *Candidatus Soleaferrea* | | | | | |
| IVW | 152 | 42.4 | -0.239 (-0.400 to -0.079) | 0.003 | 0.244 |
| MR RAPS |  |  | -0.248 (-0.418 to -0.079) | 0.004 |  |
| Weighted Median |  |  | -0.253 (-0.485 to -0.021) | 0.033 |  |
| Weighted Mode |  |  | 0.228 (-0.426 to 0.883) | 0.491 |  |
| MR Egger |  |  | -0.772 (-1.613 to 0.068) | 0.071 |  |
| Intelligence on genus *Lachnospira* | | | | | |
| IVW | 155 | 42.2 | 0.144 (0.047 to 0.242) | 0.004 | 0.197 |
| MR RAPS |  |  | 0.151 (0.048 to 0.254) | 0.004 |  |
| Weighted Median |  |  | 0.144 (7.56×10^-4^ to 0.286) | 0.049 |  |
| Weighted Mode |  |  | 0.396 (-0.010 to 0.802) | 0.056 |  |
| MR Egger |  |  | -0.096 (-0.608 to 0.415) | 0.711 |  |
| Intelligence on genus family *Defluviitaleaceae* | | | | | |
| IVW | 155 | 41.8 | 0.188 (0.050 to 0.326) | 0.008 | 0.326 |
| MR RAPS |  |  | 0.190 (0.044 to 0.336) | 0.011 |  |
| Weighted Median |  |  | 0.236 (0.042 to 0.430) | 0.017 |  |
| Weighted Mode |  |  | 0.214 (-0.361 to 0.789) | 0.464 |  |
| MR Egger |  |  | 0.935 (0.196 to 1.673) | 0.013 |  |
| Intelligence on genus *Anaerotruncus* | | | | | |
| IVW | 157 | 42.3 | 0.128 (0.032 to 0.225) | 0.009 | 0.327 |
| MR RAPS |  |  | 0.141 (0.039 to 0.243) | 0.007 |  |
| Weighted Median |  |  | 0.142 (-0.006 to 0.289) | 0.059 |  |
| Weighted Mode |  |  | 0.484 (0.074 to 0.894) | 0.021 |  |
| MR Egger |  |  | 0.317 (-0.185 to 0.819) | 0.214 |  |
| Intelligence on genus *Eubacterium eligens group* | | | | | |
| IVW | 161 | 42.2 | 0.131 (0.030 to 0.232) | 0.011 | 0.337 |
| MR RAPS |  |  | 0.125 (0.018 to 0.231) | 0.022 |  |
| Weighted Median |  |  | 0.106 (-0.044 to 0.255) | 0.167 |  |
| Weighted Mode |  |  | 0.009 (-0.360 to 0.379) | 0.960 |  |
| MR Egger |  |  | 0.481 (-0.053 to 1.014) | 0.077 |  |
| Intelligence on genus *Coprococcus2* | | | | | |
| IVW | 159 | 42 | 0.141 (0.026 to 0.257) | 0.017 | 0.44 |
| MR RAPS |  |  | 0.143 (0.021 to 0.265) | 0.022 |  |
| Weighted Median |  |  | 0.106 (-0.053 to 0.264) | 0.190 |  |
| Weighted Mode |  |  | -0.079 (-0.588 to 0.430) | 0.760 |  |
| MR Egger |  |  | 0.172 (-0.439 to 0.783) | 0.579 |  |
| Intelligence on genus *Ruminococcus2* | | | | | |
| IVW | 150 | 41.6 | -0.117 (-0.220 to -0.013) | 0.027 | 0.635 |
| MR RAPS |  |  | -0.124 (-0.233 to -0.015) | 0.026 |  |
| Weighted Median |  |  | -0.162 (-0.308 to -0.017) | 0.029 |  |
| Weighted Mode |  |  | -0.432 (-0.876 to 0.012) | 0.056 |  |
| MR Egger |  |  | -0.338 (-0.885 to 0.210) | 0.225 |  |
| Intelligence on genus *Roseburia* | | | | | |
| IVW | 152 | 42.2 | 0.104 (0.009 to 0.199) | 0.031 | 0.658 |
| MR RAPS |  |  | 0.107 (0.007 to 0.207) | 0.036 |  |
| Weighted Median |  |  | 0.034 (-0.101 to 0.169) | 0.624 |  |
| Weighted Mode |  |  | -0.143 (-0.627 to 0.342) | 0.562 |  |
| MR Egger |  |  | 0.009 (-0.491 to 0.510) | 0.971 |  |
| Intelligence on family *Bacteroidales S24 7group* | | | | | |
| IVW | 157 | 42.3 | 0.143 (0.003 to 0.282) | 0.045 | 0.868 |
| MR RAPS |  |  | 0.145 (-0.002 to 0.292) | 0.053 |  |
| Weighted Median |  |  | 0.097 (-0.101 to 0.294) | 0.338 |  |
| Weighted Mode |  |  | 0.088 (-0.412 to 0.588) | 0.728 |  |
| MR Egger |  |  | 0.237 (-0.493 to 0.967) | 0.522 |  |
| Intelligence on unknown genus id.1000005479 | | | | | |
| IVW | 157 | 42.3 | 0.143 (0.003 to 0.282) | 0.045 | 0.796 |
| MR RAPS |  |  | 0.145 (-0.002 to 0.292) | 0.053 |  |
| Weighted Median |  |  | 0.097 (-0.105 to 0.298) | 0.348 |  |
| Weighted Mode |  |  | 0.088 (-0.411 to 0.587) | 0.728 |  |
| MR Egger |  |  | 0.237 (-0.493 to 0.967) | 0.522 |  |
| Intelligence on genus *Prevotella9* | | | | | |
| IVW | 158 | 41.7 | -0.126 (-0.250 to -0.001) | 0.047 | 0.769 |
| MR RAPS |  |  | -0.128 (-0.259 to 0.002) | 0.054 |  |
| Weighted Median |  |  | -0.050 (-0.234 to 0.134) | 0.595 |  |
| Weighted Mode |  |  | 0.118 (-0.409 to 0.644) | 0.660 |  |
| MR Egger |  |  | -0.496 (-1.147 to 0.155) | 0.134 |  |
| Intelligence on genus *Lachnospiraceae NC2004 group* | | | | | |
| IVW | 156 | 42.1 | 0.159 ( 2.36×10^-4^ to 0.317) | 0.050 | 0.748 |
| MR RAPS |  |  | 0.153 (-0.013 to 0.320) | 0.071 |  |
| Weighted Median |  |  | 0.061 (-0.173 to 0.295) | 0.611 |  |
| Weighted Mode |  |  | -0.374 (-0.956 to 0.208) | 0.206 |  |
| MR Egger |  |  | -0.094 (-0.925 to 0.737) | 0.824 |  |
| Intelligence on genus *Alloprevotella* | | | | | |
| IVW | 105 | 43.1 | -0.276 (-0.552 to -4.93×10^-5^) | 0.050 | 0.703 |
| MR RAPS |  |  | -0.277 (-0.568 to 0.014) | 0.062 |  |
| Weighted Median |  |  | -0.193 (-0.593 to 0.207) | 0.345 |  |
| Weighted Mode |  |  | -0.081 (-1.124 to 0.962) | 0.879 |  |
| MR Egger |  |  | -0.237 (-1.758 to 1.284) | 0.758 |  |

MR: Mendelian Randomization; IVW: inverse variance–weighted; RAPS: robust adjusted profile score; SNP: single nucleotide polymorphism; CI: confidence intervals. P-values from the IVW MR test were adjusted using Benjamini–Hochberg FDR correction.

**Supplementary Table 11. Summary information on brain volume for the 12 genome-wide significant SNPs associated with genus *Oxalobacter***

| **SNP** | **A1** | **A2** | **EAF** | **Beta** | **SE** | **N** | ***P*-value** |
| --- | --- | --- | --- | --- | --- | --- | --- |
| rs4428215 | A | G | 0.774 | -0.004 | 0.008 | 47282 | 0.622 |
| rs736744 | T | C | 0.604 | -0.003 | 0.007 | 47179 | 0.600 |
| rs6000536 | T | C | 0.801 | -0.002 | 0.008 | 46649 | 0.777 |
| rs36057338 | T | G | 0.932 | 0.000 | 0.013 | 47284 | 0.984 |
| rs6071435 | A | T | 0.631 | 0.012 | 0.008 | 35900 | 0.117 |
| rs12002250 | A | C | 0.055 | 0.002 | 0.014 | 47196 | 0.913 |
| rs1569853 | T | C | 0.154 | 0.004 | 0.009 | 47305 | 0.670 |
| rs11108500 | A | G | 0.067 | 0.015 | 0.013 | 47316 | 0.242 |
| rs10464997 | A | G | 0.868 | -0.003 | 0.010 | 47060 | 0.762 |
| rs111966731^a^ | T | C | 0.061 | -0.040 | 0.014 | 47303 | 0.003 |
| rs6993398 | A | G | 0.859 | 0.005 | 0.009 | 47268 | 0.575 |
| rs3862635 | T | C | 0.936 | 0.011 | 0.013 | 47276 | 0.394 |

SNP: single nucleotide polymorphism; A1: effect allele; A2: baseline allele; EAF: effect allele frequency; SE: standard error. ^a^ SNPs were removed in the MR analysis during the heterogeneity test via RadialMR.

**Supplementary Table 12. Summary information on brain volume for the 19 genome-wide significant SNPs associated with genus *Fusicatenibacter***

| **SNP ^a^** | **A1** | **A2** | **EAF** | **Beta** | **SE** | **N** | ***P*-value** |
| --- | --- | --- | --- | --- | --- | --- | --- |
| rs4378146 | A | C | 0.251 | -0.013 | 0.008 | 47136 | 0.088 |
| rs206581 | A | G | 0.223 | 0.006 | 0.008 | 46855 | 0.459 |
| rs62187631 | T | C | 0.152 | -0.008 | 0.009 | 44786 | 0.406 |
| rs2025938 | A | G | 0.919 | 0.006 | 0.012 | 47197 | 0.639 |
| rs3303 | T | C | 0.084 | -0.005 | 0.012 | 46998 | 0.685 |
| rs2039204 ^b^ | A | T | 0.515 | 0.012 | 0.007 | 35741 | 0.118 |
| rs8028026 | A | G | 0.112 | -0.017 | 0.010 | 47235 | 0.094 |
| rs1864685 | A | C | 0.477 | -0.009 | 0.007 | 47209 | 0.169 |
| rs792108 | T | C | 0.356 | 0.002 | 0.007 | 47316 | 0.748 |
| rs60254196 | A | G | 0.514 | 0.004 | 0.007 | 46963 | 0.499 |
| rs9905659 | A | G | 0.823 | -0.004 | 0.009 | 46820 | 0.620 |
| rs8063430 | T | C | 0.055 | -0.025 | 0.014 | 47136 | 0.076 |
| rs6515626 | A | G | 0.957 | -0.022 | 0.016 | 47316 | 0.181 |
| rs10439674 | A | G | 0.202 | -0.003 | 0.008 | 47200 | 0.674 |
| rs167879 | T | C | 0.827 | -0.004 | 0.009 | 47276 | 0.633 |
| rs73103914 | A | G | 0.211 | -0.015 | 0.008 | 47183 | 0.069 |

SNP: single nucleotide polymorphism; A1: effect allele; A2: baseline allele; EAF: effect allele frequency; SE: standard error. ^a^ rs2132128, rs62353480 and rs704418 were not available in the outcome GWAS, and LDlink didn’t detect closely related SNP (r^2^ > 0.8) associated with the *genus Fusicatenibacter* (*P* < 1×10^-5^) and available in the outcome GWAS. Thus, these three SNPs were removed in the MR analysis. ^b^ Palindromic SNPs with intermediate allele frequencies (>0.42) were removed.

**Supplementary Table 13. Pleiotropy assessment results for the mediation analysis**

| **Method** | ***Oxalobacter* on brain volume** | ***Fusicatenibacter* on brain volume** | **Brain volume on intelligence** |
| --- | --- | --- | --- |
| MR Egger intercept | 0.013 | -0.011 | 0.008 |
| MR Egger intercept 95%CI | -0.013 to 0.038 | -0.027 to 0.005 | -0.003 to 0.019 |
| MR Egger intercept *P* | 0.363 | 0.215 | 0.195 |
| MRPRESSO Global Test RSSobs | 6.29 | 15.00 | 7.87 |
| MRPRESSO Global Test *P* | 0.857 | 0.577 | 0.763 |
| Cochran’s Q | 5.30 | 12.14 | 6.38 |
| Cochran’s Q *P* | 0.870 | 0.595 | 0.701 |
| Rucker’s Qʹ | 4.38 | 10.44 | 4.38 |
| Rucker’s Qʹ *P* | 0.884 | 0.658 | 0.822 |
| Q-Q' | 0.92 | 1.70 | 2.00 |
| Q-Q' *P* | 0.338 | 0.192 | 0.157 |

**Supplementary Table 14. Summary information on brain volume used as genetic instruments for the Mendelian randomization analyses**

| **SNP** | **A1** | **A2** | **EAF** | **Beta** | **SE** | **N** | ***P*-value** | **R^2^ (%)** | **F-statistics** |
| --- | --- | --- | --- | --- | --- | --- | --- | --- | --- |
| rs10927041 | T | C | 0.815 | -0.057 | 0.008 | 47280 | 8.16E-12 | 0.099 | 46.7 |
| rs7608881 | C | G | 0.522 | 0.045 | 0.007 | 35939 | 2.02E-09 | 0.100 | 36.0 |
| rs288326 | A | G | 0.119 | 0.070 | 0.010 | 47316 | 3.99E-12 | 0.102 | 48.1 |
| rs34514405 | A | G | 0.849 | -0.060 | 0.009 | 47298 | 2.94E-11 | 0.093 | 44.2 |
| rs3134953 | A | G | 0.857 | -0.059 | 0.010 | 45055 | 5.01E-10 | 0.086 | 38.7 |
| rs2764264 | T | C | 0.697 | 0.066 | 0.007 | 47085 | 7.92E-21 | 0.186 | 87.6 |
| rs4273712 | A | G | 0.733 | -0.069 | 0.007 | 47316 | 9.68E-21 | 0.184 | 87.2 |
| rs151057105 | T | C | 0.092 | 0.071 | 0.011 | 47184 | 2.54E-10 | 0.085 | 40.0 |
| rs42035 | A | G | 0.756 | 0.051 | 0.008 | 47171 | 2.10E-11 | 0.095 | 44.9 |
| rs478839 | A | G | 0.609 | 0.036 | 0.007 | 47185 | 4.55E-08 | 0.063 | 29.9 |
| rs1628768 | T | C | 0.766 | -0.052 | 0.008 | 47190 | 9.71E-12 | 0.098 | 46.4 |
| rs3217870 | T | C | 0.607 | -0.040 | 0.007 | 47133 | 3.02E-09 | 0.075 | 35.2 |
| rs2066827 | T | G | 0.767 | 0.050 | 0.008 | 43580 | 4.02E-10 | 0.090 | 39.1 |
| rs7970368 | T | G | 0.823 | -0.055 | 0.009 | 47260 | 1.42E-10 | 0.087 | 41.1 |
| rs7297175 | T | C | 0.432 | 0.045 | 0.007 | 47260 | 7.98E-12 | 0.099 | 46.8 |
| rs7306710 | T | C | 0.479 | 0.048 | 0.007 | 46883 | 1.74E-13 | 0.116 | 54.3 |
| rs28675824 | C | G | 0.205 | 0.054 | 0.009 | 35901 | 3.70E-09 | 0.097 | 34.8 |
| rs62057149 | A | G | 0.775 | 0.078 | 0.008 | 47038 | 2.72E-23 | 0.210 | 98.9 |

SNP: single nucleotide polymorphism; A1: effect allele; A2: baseline allele; EAF: effect allele frequency; SE: standard error; R^2^: Explained incremental variance in the phenotype.

**Supplementary Table 15. Summary information on intelligence for the 18 genome-wide significant SNPs associated with brain volume**

| **SNP ^a^** | **A1** | **A2** | **EAF** | **Beta** | **SE** | **N** | ***P*-value** |
| --- | --- | --- | --- | --- | --- | --- | --- |
| rs10927041 | T | C | 0.804 | -0.007 | 0.003 | 268494 | 0.051 |
| rs7608881^a^ | G | A | 0.444 | -0.010 | 0.003 | 266441 | 2.90E-04 |
| rs288326 | G | A | 0.880 | -0.006 | 0.004 | 267504 | 0.175 |
| rs34514405^b^ | A | G | 0.850 | -0.019 | 0.004 | 265696 | 5.50E-07 |
| rs2764264^b^ | C | T | 0.345 | -0.023 | 0.003 | 262121 | 3.76E-15 |
| rs4273712^b^ | A | G | 0.726 | -0.014 | 0.003 | 269356 | 2.44E-06 |
| rs151057105 | C | T | 0.900 | -0.014 | 0.005 | 261376 | 0.002 |
| rs42035 | A | G | 0.751 | 0.006 | 0.003 | 263182 | 0.058 |
| rs478839^c^ | A | G | 0.588 | 0.001 | 0.003 | 266789 | 0.831 |
| rs1628768 | T | C | 0.753 | -0.007 | 0.003 | 263526 | 0.029 |
| rs3217870 | C | T | 0.374 | 0.007 | 0.003 | 258063 | 0.018 |
| rs2066827 | T | G | 0.757 | 0.011 | 0.003 | 242741 | 0.002 |
| rs7970368 | T | G | 0.827 | -0.011 | 0.004 | 265491 | 0.002 |
| rs7297175^b^ | T | C | 0.417 | 0.014 | 0.003 | 264698 | 2.48E-07 |
| rs7306710 | T | C | 0.458 | 0.008 | 0.003 | 259699 | 0.004 |
| rs28675824^b^ | G | C | 0.789 | -0.016 | 0.003 | 265137 | 2.68E-06 |
| rs62057149^b^ | A | G | 0.794 | 0.016 | 0.003 | 263412 | 1.61E-06 |

SNP: single nucleotide polymorphism; A1: effect allele; A2: baseline allele; EAF: effect allele frequency; SE: standard error. ^a^ rs3134953 was not available in the outcome GWAS, and LDlink didn’t detect closely related SNP (r^2^ > 0.8) associated with brain volume (*P* < 5×10^-8^) and available in the outcome GWAS. Thus, rs3134953 was removed in the MR analysis. rs7608881 was also not available in the outcome GWAS, we replaced it with closely related SNP rs7608892. ^b^ SNPs were removed from the analysis due to a strong association with the outcome (*P* < 1×10^-5^). ^c^ SNPs were removed in the MR analysis during the heterogeneity test via RadialMR.

**Supplementary Figure 1. Leave-one-out analyses for SNPs associated with genus *Oxalobacter* and genus *Fusicatenibacter* on intelligence**

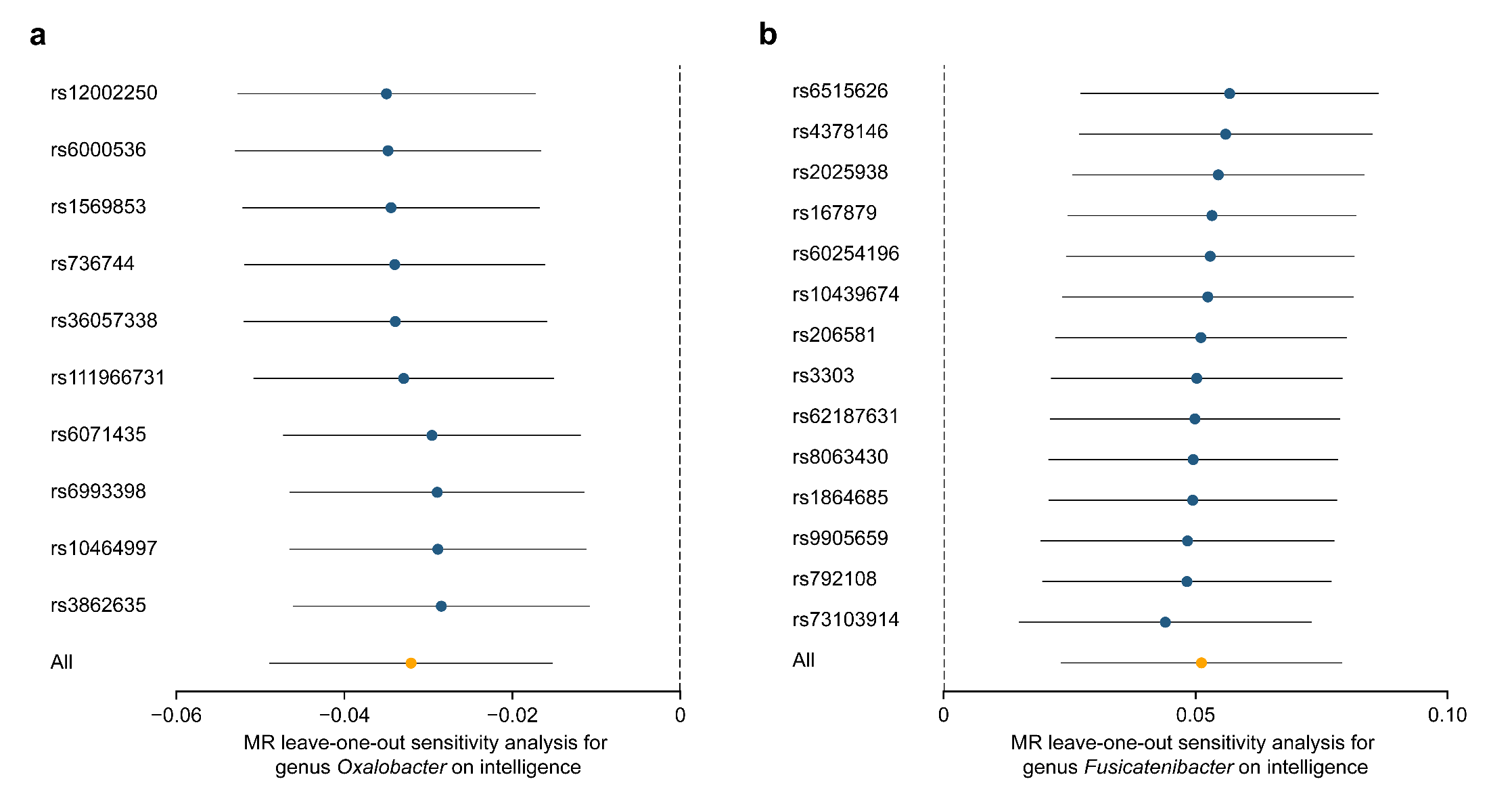

The leave-one-SNP out analysis was conducted to assess the influence of individual variants of genus *Oxalobacter* and genus *Fusicatenibacter* on intelligence. Error bars represent 95% confidence intervals.

**Supplementary Figure 2. Mendelian randomization plots for the relationship of intelligence with genus *Oxalobacter* and genus *Fusicatenibacter***

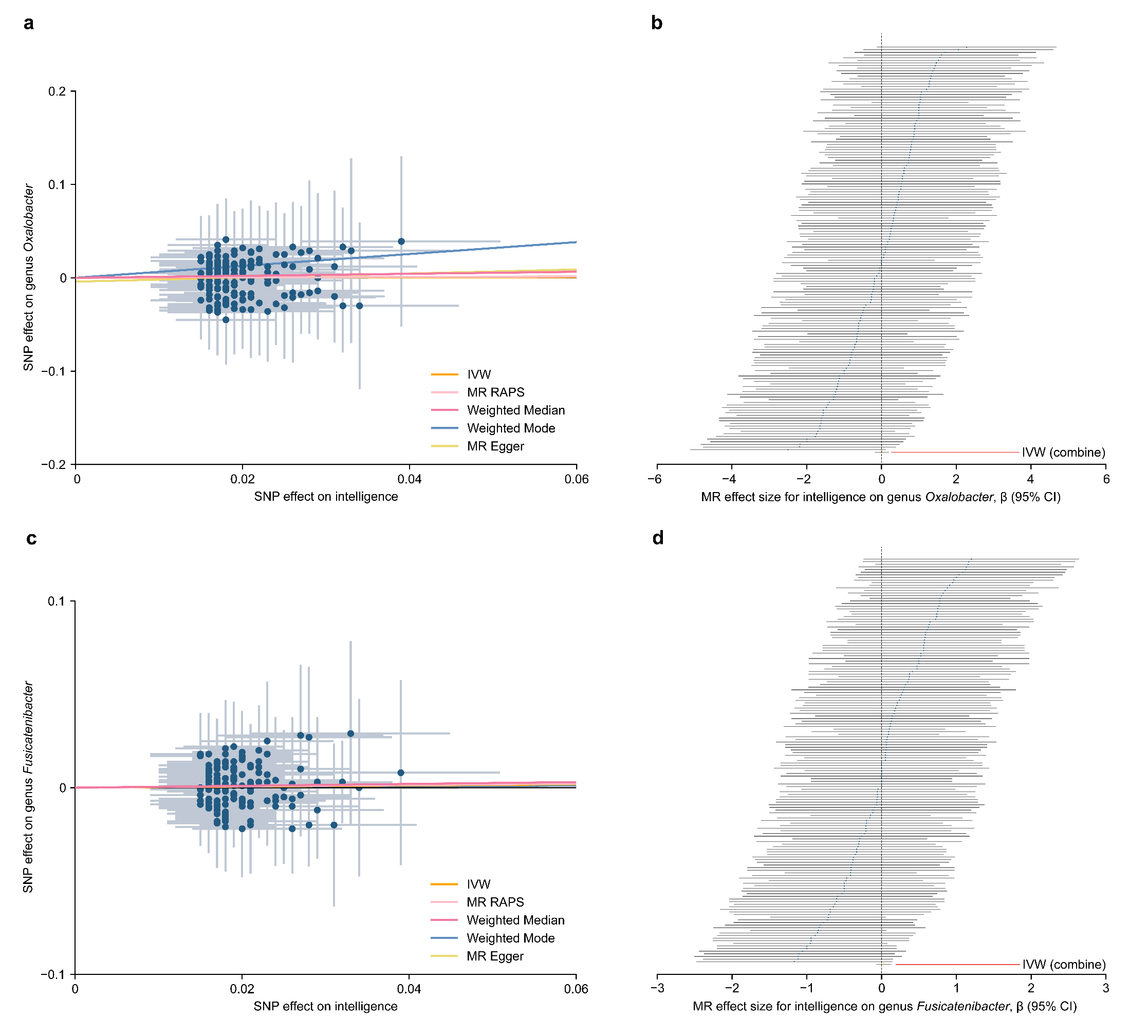

Scatter plot of SNP effects on intelligence versus genus *Oxalobacter* (a), and intelligence versus genus *Fusicatenibacter* (c), with the slope of each line corresponding to the estimated MR effect per method. The data are expressed as raw β values with 95% CIs. Forest plot of individual and combined SNP MR-estimated effect sizes for intelligence on genus *Oxalobacter* (b), and intelligence on genus *Fusicatenibacter* (d). Error bars represent 95% confidence intervals.

**Supplementary Figure 3. Leave-one-out analyses for SNPs associated with intelligence on genus *Oxalobacter* and genus *Fusicatenibacter***

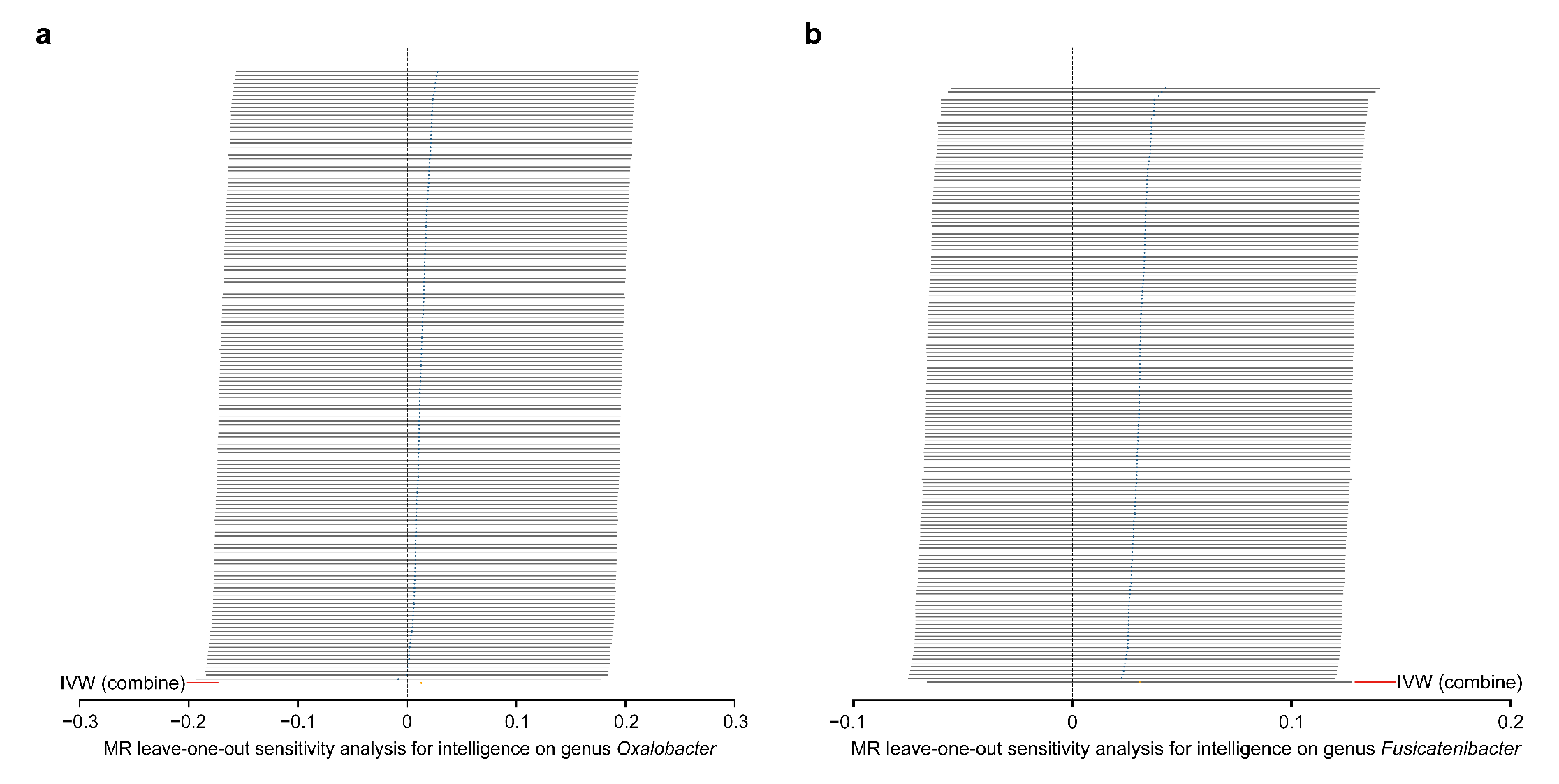

The leave-one-SNP out analysis was conducted to assess the influence of individual variants of intelligence on genus *Oxalobacter* and genus *Fusicatenibacter*. Error bars represent 95% confidence intervals.

**Supplementary Figure 4. Mendelian randomization plots for the relationship of genus *Oxalobacter* and genus *Fusicatenibacter* with brain volume**

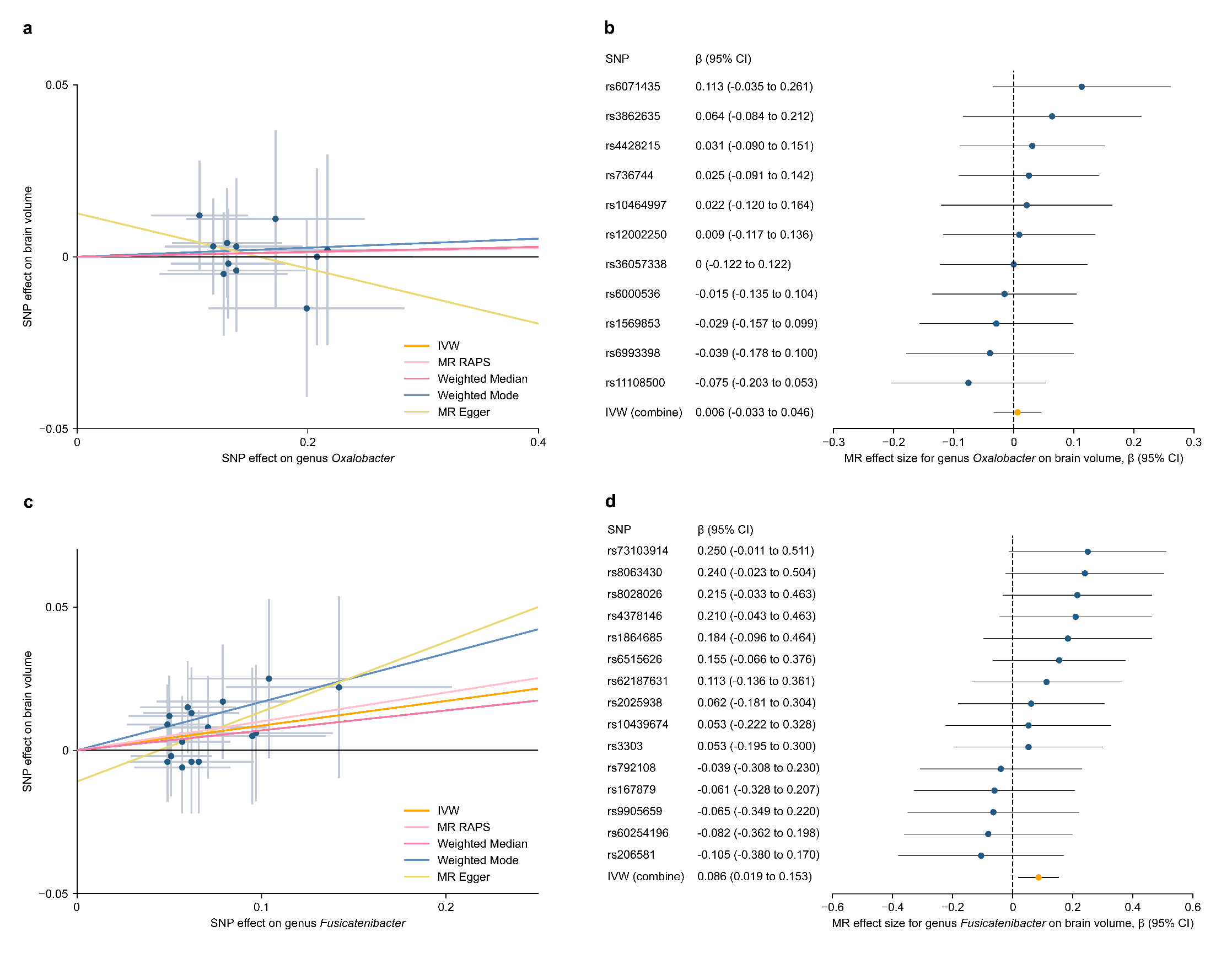

Scatter plot of SNP effects on genus *Oxalobacter* versus brain volume (a), and genus *Fusicatenibacter* versus brain volume (c), with the slope of each line corresponding to the estimated MR effect per method. The data are expressed as raw β values with 95% CIs. Forest plot of individual and combined SNP MR-estimated effect sizes for genus *Oxalobacter* on brain volume (b), and genus *Fusicatenibacter* on brain volume (d). Error bars represent 95% confidence intervals.

**Supplementary Figure 5. Leave-one-out analyses for SNPs associated with genus *Oxalobacter* and genus *Fusicatenibacter* on brain volume**

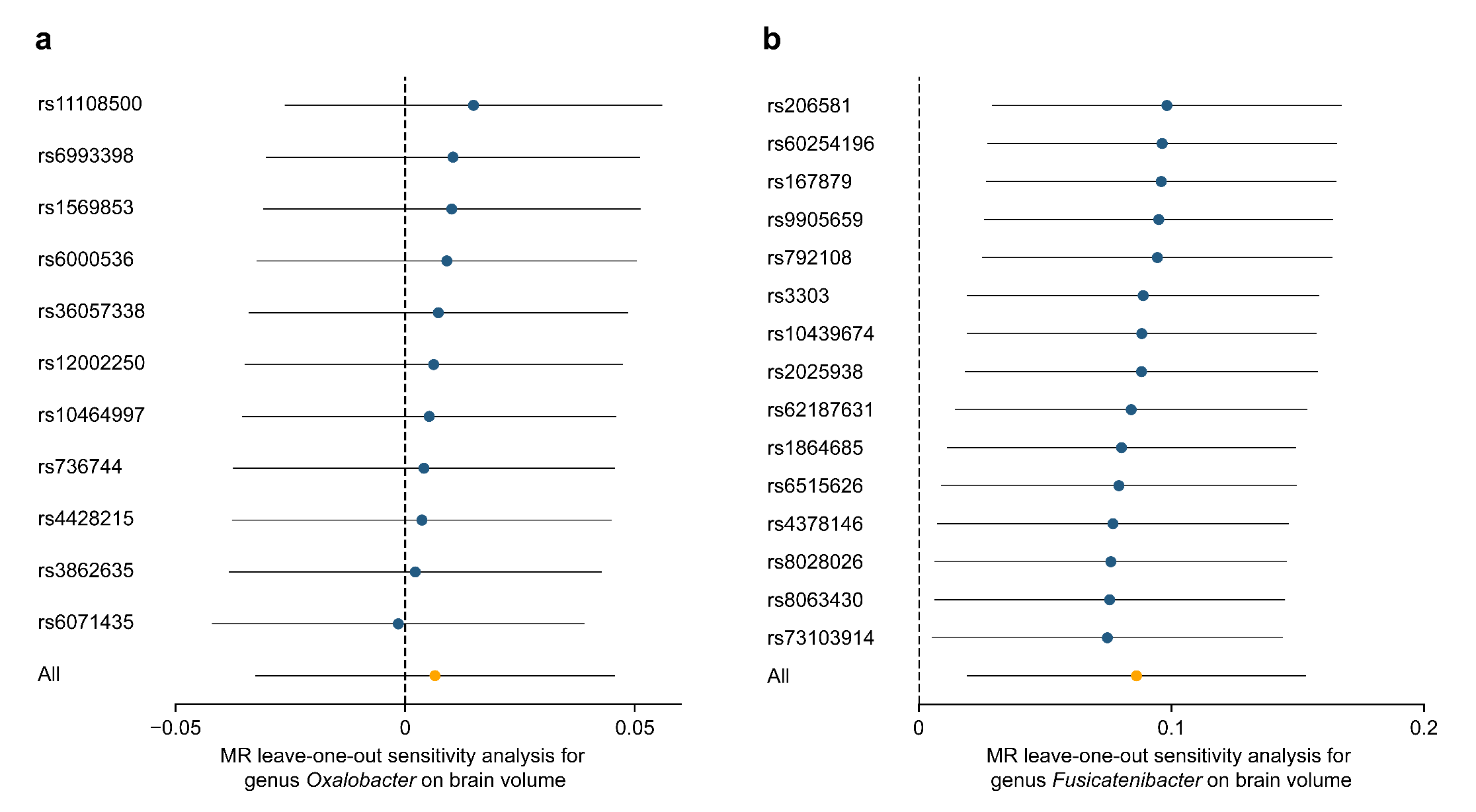

The leave-one-SNP out analysis was conducted to assess the influence of individual variants of genus *Oxalobacter* and genus *Fusicatenibacter* on brain volume. Error bars represent 95% confidence intervals.

**Supplementary Figure 6. Mendelian randomization plots for the relationship of brain volume with intelligence**

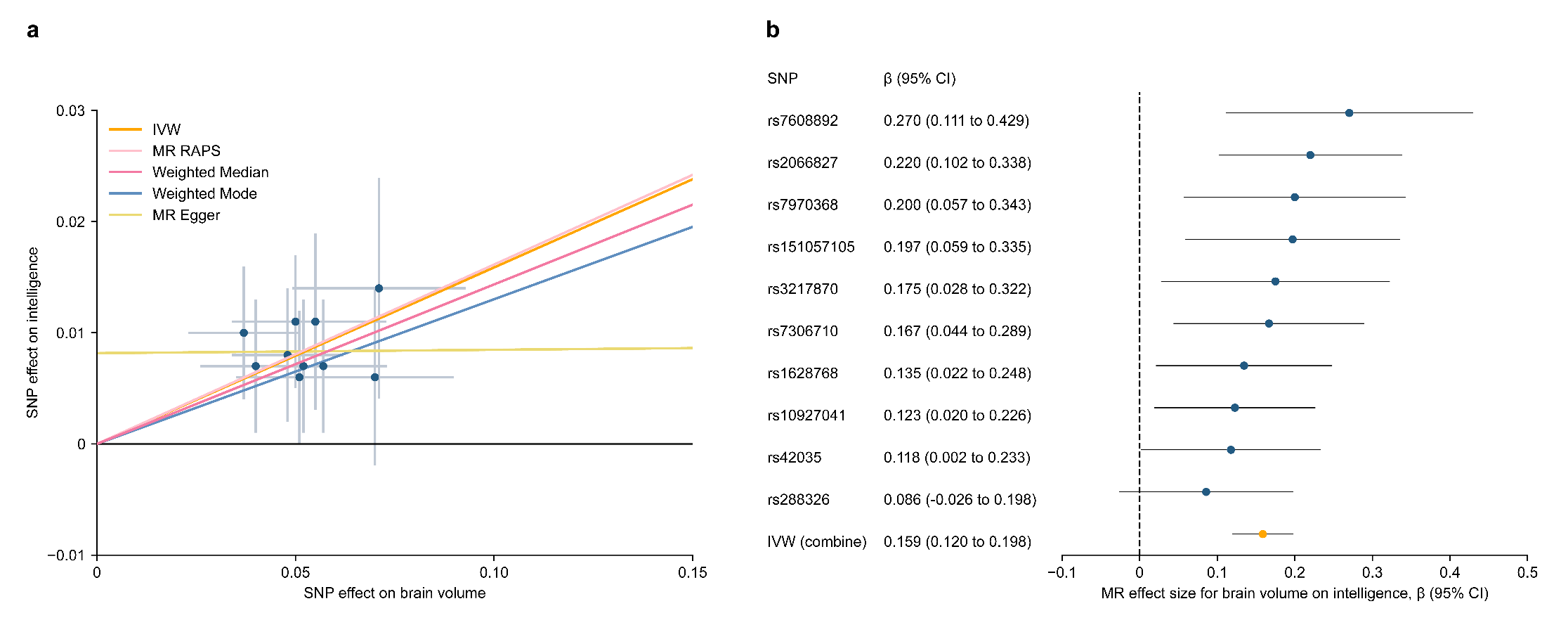

Scatter plot of SNP effects on brain volume versus intelligence (a), with the slope of each line corresponding to the estimated MR effect per method. The data are expressed as raw β values with 95% CIs. Forest plot of individual and combined SNP MR-estimated effect sizes for brain volume on intelligence (b). Error bars represent 95% confidence intervals.

**Supplementary Figure 7. Leave-one-out analyses for SNPs associated with brain volume on intelligence**

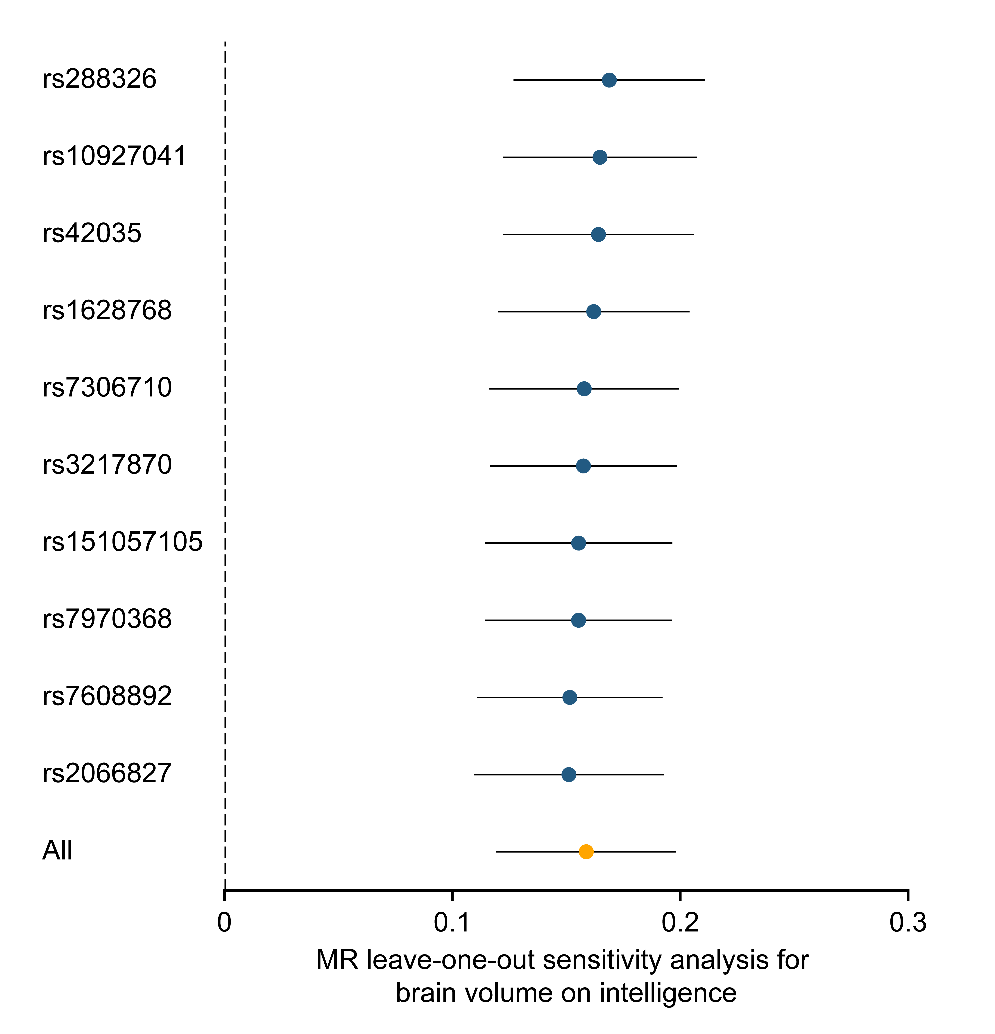

The leave-one-SNP out analysis was conducted to assess the influence of individual variants of brain volume on intelligence. Error bars represent 95% confidence intervals.
